# Evidence for Behavioral Autorepression in Covid-19 Epidemiological Dynamics

**DOI:** 10.1101/2024.06.07.24308626

**Authors:** Daniel D. Lewis, Michael Pablo, Xinyue Chen, Michael L. Simpson, Leor Weinberger

## Abstract

It has long been hypothesized that behavioral reactions to epidemic severity autoregulate infection dynamics, for example when susceptible individuals self-sequester based on perceived levels of circulating disease. However, evidence for such ‘behavioral autorepression’ has remained elusive, and its presence could significantly affect epidemic forecasting and interventions. Here, we analyzed early COVID-19 dynamics at 708 locations over three epidemiological scales (96 countries, 50 US states, and 562 US counties). Signatures of behavioral autorepression were identified through: (i) a counterintuitive mobility-death correlation, (ii) fluctuation-magnitude analysis, and (iii) dynamics of SARS-CoV-2 infection waves. These data enabled calculation of the average behavioral-autorepression strength (i.e., negative feedback ‘gain’) across different populations. Surprisingly, incorporating behavioral autorepression into conventional models was required to accurately forecast COVID-19 mortality. Models also predicted that the strength of behavioral autorepression has the potential to alter the efficacy of non-pharmaceutical interventions. Overall, these results provide evidence for the long-hypothesized existence of behavioral autorepression, which could improve epidemic forecasting and enable more effective application of non-pharmaceutical interventions during future epidemics.

**Significance:** Challenges with epidemiological forecasting during the COVID-19 pandemic suggested gaps in underlying model architecture. One long-held hypothesis, typically omitted from conventional models due to lack of empirical evidence, is that human behaviors lead to intrinsic negative autoregulation of epidemics (termed ‘behavioral autorepression’). This omission substantially alters model forecasts. Here, we provide independent lines of evidence for behavioral autorepression during the COVID-19 pandemic, demonstrate that it is sufficient to explain counterintuitive data on ‘shutdowns’, and provides a mechanistic explanation of why early shutdowns were more effective than delayed, high-intensity shutdowns. We empirically measure autorepression strength, and show that incorporating autorepression dramatically improves epidemiological forecasting. The autorepression phenomenon suggests that tailoring interventions to specific populations may be warranted.

## INTRODUCTION

The dynamics of communicable diseases have historically been analyzed using Susceptible-Infective-Removed (SIR) models (1), wherein the rate of infection spread is mediated by density-dependent contact between the susceptible and infective populations, generating an inherent positive-feedback or auto-stimulatory loop. However, it has long been hypothesized that these infection rates can be buffered by human apprehension, thereby generating a form of negative feedback or autorepression. This phenomenon of “behavioral autorepression” postulates that, based on the size of the infective population, susceptible individuals self-sequester during an epidemic to reduce the effective contact rate and interrupt the spread of disease.

Behavioral autorepression was first proposed in mathematical models of a cholera outbreak in the summer of 1973 centered around the Italian town of Bari on the Mediterranean coast (2, 3). Epidemiological data showed that daily cholera infections plateaued, rather than continuing to exponentially grow, and models argued this was due to reductions in contact rates as a result of human apprehension. The resulting behavioral autorepression models exhibited plateaus in daily infections through saturation of the contact rate, in contrast with canonical SIR models which predicted exponential rises and a peak in daily infections, followed by a steady drop (1).

Despite the long history of the behavioral autorepression hypothesis, other epidemiological mechanisms have been proposed as alternate explanations of infection-rate saturation. For example, lockdown-limited transmission (4) can mimic the effects of autorepression, and depletion of high-risk individuals (5–7) can also generate comparable saturation of infection rates. Consequently, the role of behavioral autorepression and its relative contribution to population-scale disease transmission remain unclear.

The lack of clarity surrounding behavioral autorepression has significant implications for epidemiological forecasts. In principle, behavioral autorepression would substantially reduce predicted infections and deaths in SIR-like models, potentially leading to improved forecasting of epidemic dynamics. Additionally, theoretical models have shown behavioral autorepression to be capable of generating multiple waves of infection (8, 9).

This potential of behavioral autorepression to influence disease dynamics catalyzed substantial modeling efforts to explore its epidemiologic impacts. Hypotheses suggested that autorepression may be a form of natural infection mitigation, since conventional policy interventions (a.k.a., non-pharmaceutical interventions) are notoriously challenging to maintain due to fatigue and poor adherence (10). Simulation-based approaches were used to examine how changes in the mathematical formulation of behavioral autorepression affect the sensitivity of epidemic dynamics to parameter changes (11, 12). Studies also modeled how changes in media coverage influenced changes in population-level psychology and fear to affect the propagation of cases numbers (9, 13, 14).

Despite the potential importance of behavioral autorepression, previous disease outbreaks that may have manifested autorepression dynamics occurred during eras when consistent population-level behavioral metrics were largely unavailable (15). In contrast, the level of data collection during the COVID-19 pandemic was unprecedented. For example, modern improvements in epidemiological data recording practices were employed (16), and there was broad availability of mobile-phone geolocation data (17), which can quantify regional, population-level behavior at multiple geographic scales (18, 19). Thus, we hypothesized these datasets could enable a unique empirical analysis of an acute outbreak to quantify how human responses (e.g., behavioral autorepression) influence contact rates and disease transmission.

Here, we examined various COVID-19 epidemiological data for signatures of autorepression (i.e., negative feedback) using population mobility data as a correlate of the contact rate. Analysis of the early epidemic (i.e., spring 2020) showed a counterintuitive time-dependent inversion of the correlation between COVID-19 deaths and population mobility across different epidemiological scales (96 countries, 50 states, and 562 counties). This inversion was consistent with autorepression but not with alternate epidemiological mechanisms. Independent lines of evidence for behavioral autorepression were established by assaying for signatures of autorepression in the dynamics of COVID-19 infection. Fluctuation analysis of daily infection counts showed direct evidence of significantly altered epidemic feedback strength. Analysis of the first wave of COVID-19 infections showed that the timing of waves was positively correlated with wave intensity, consistent with variation in autorepression delay. Based on these data indicating the existence of behavioral autorepression, we applied behavioral autorepression to SIR-type models and found that autorepression dramatically improved the mortality forecasting of these models.

## RESULTS

### Epidemiological data show unexpected inversion of the mobility-death correlation

To search for potential signatures of population-level behavioral autorepression, we examined temporal changes in population-level mobility (a correlate of contact rate) early in the pandemic on a region-by-region basis, relative to longitudinal COVID-19 mortality data. To avoid infection underreporting, we focused on confirmed COVID-19 deaths as the measure for disease transmission (20), and to quantify changes in population-level behavior, we used mobile phone geolocation data from Google’s mobility dataset (21), where location data is broken down into occupancy of different location classes: (i) retail & recreation occupancy, (ii) grocery & pharmacy occupancy, (iii) park occupancy, (iv) transit occupancy, (v) workplace occupancy, and (vi) residential occupancy. We hypothesized that residential occupancy likely represents the inverse aggregate of all the other mobility measures, so our initial analyses focused on residential occupancy as a surrogate for occupancy changes in multiple categories of public spaces (i–v), whereas subsequent analyses directly analyzed these other mobility measures.

Using linear regression, we examined two aspects of occupancy: (i) the change in occupancy from baseline after a threshold of ten deaths had been crossed (termed “*initial change in residential occupancy*”); and (ii) the maximum change in residential occupancy during the first wave of COVID-19 infection (termed “*maximum change in residential occupancy”*), which typically occurred around thirty days after the ten-death threshold was crossed in a region. To examine the relationship between mobility and mortality during the first wave of COVID-19, both occupancy metrics (i.e., initial and max changes) were compared to COVID-19 deaths during the first wave on a region-to-region basis (**Fig. 1**). Regression analysis was performed at three different geographic scales––from coarse-grained to finer-grained: (i) international (96 countries); (ii) provincial (50 US states); and (iii) regional (562 US counties). As expected, regression analysis of the coarse-grained data of 96 countries revealed that initial changes in residential occupancy were negatively correlated with deaths per capita (**Fig. 1A**, left). However, the maximum change in residential occupancy appeared counterintuitively inverted, showing a positive correlation with regional mortality (**Fig. 1A**, right). To ensure that this observed inversion of the correlation was not an artifact of coarse-grained geographical analysis, we repeated the analysis at the provincial and regional level. These finer-grained analyses of states and counties in the United States showed very similar inversion of the mobility-death correlation (**Fig. 1B–C**).

**Fig. 1:**
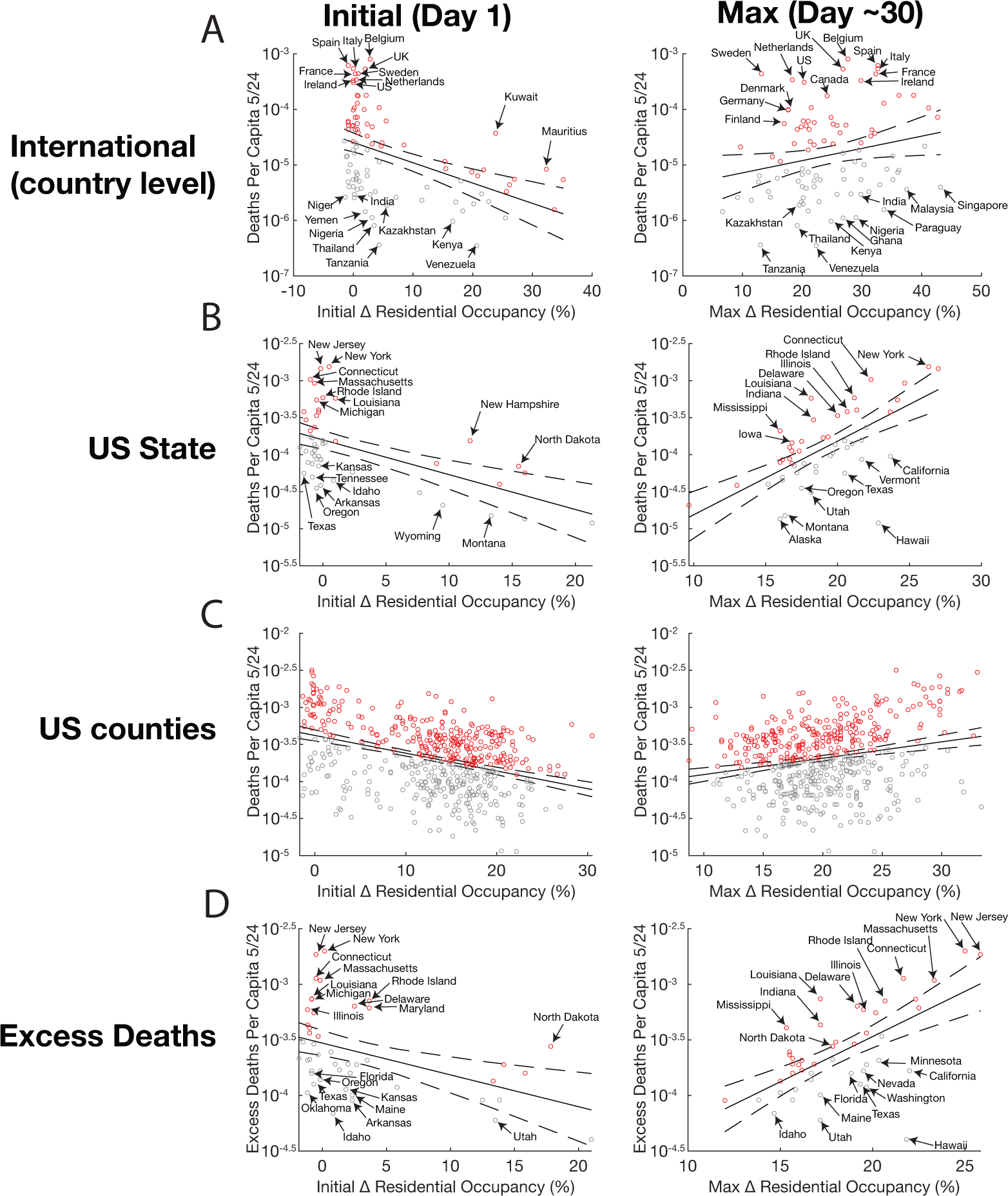
Temporal inversion of the mobility-death correlation early in the COVID-19 outbreak. COVID-19 deaths per capita vs. population mobility (as measured by residential occupancy) from mobile-phone data. Initial and max Δ occupancy is defined as deviation from baseline occupancy at the start of the epidemic, defined as 10 deaths (left, day 1) or the maximum deviation from baseline occupancy (right, day ∼30), respectively; deaths per capita are totaled until May 24^th^, 2020. Solid lines are the linear regression, dashed lines are 95% confidence intervals. Red points represent regions that had higher death than predicted by the regression. Grey points represent regions that had lower death than predicted by the regression. **(A**) Data of international of COVID-19 deaths vs. initial change in residential occupancy (Left; regression p-value=6.7×10^−6^) and max. increase in residential occupancy (Right; regression p-value= 3.2×10^−2^). (**B**) US-state COVID-19 deaths vs. initial change in residential occupancy (Left, regression p=2.7×10^−5^) and max. increase in residential occupancy (Right; regression p-value=1.9×10^−7^). (**C**) US county COVID-19 deaths versus initial change in residential occupancy (Left; regression p-value=8.2 × 10^−19^) and max. increase in residential occupancy (Right; regression p-value=1.9×10^−7^). (**D**) Excess deaths (US state) vs. initial change in residential occupancy (Left; regression p-value=1.7×10^−3^) and max. change in residential occupancy (Right; regression p-value=1.8×10^−6^). P-values indicate significance that the slope of the regression line is non-zero (null hypothesis slope=0).

To eliminate the possibility that incomplete death statistics (22) might generate a spurious mobility-death correlation, we repeated the regression analysis using excess deaths instead of confirmed COVID-19 deaths. Estimates of excess deaths associated with COVID-19 from the CDC were calculated by comparing mortality rates during the COVID-19 pandemic to average mortality statistics from previous years (23–25). Despite using excess deaths, the inverted correlation between initial and maximum residential occupancy persisted (**Fig. 1D**). Excess deaths also showed an inverted correlation between initial and maximum change in retail, transit, and workplace occupancy (**Fig. S1**). These results suggest that the inversion of the mobility-death correlation is not caused by an incomplete characterization of COVID-19 mortality (**Fig. 1D, Fig. S1**).

To verify that the observed inversion of the mobility-death correlation was not an artifact of the particular mobility metric used (i.e., residential occupancy), we analyzed mortality data versus the five other mobility categories. These other metrics represent occupancy of public spaces which decreased during the first wave of COVID-19 (as opposed to residential occupancy, which increased during the first wave of COVID-19). Retail & recreation, transit, and workplace occupancy all exhibited temporal inversion of the mortality-death correlation (**Fig. S2-S6**). Overall, alternate measures of mobility showed a similar temporal inversion of the mobility-death correlation.

We additionally tested that the inversion of the mobility-death correlation was retained with alternative regression methods and alternative sources for mobility data (Supp Text-Section 1). In all cases, the inversion of the mobility-death correlation was found to be a robust phenomenon that occurred across multiple geographic scales, irrespective of data source or underlying statistical assumptions (**Fig. S7-S10**).

### Minimal autorepression models explain inversion of the mobility-death correlation

Building off the prior finding that population-level mobility affects contact rate (26), these empirical data (**Fig. 1**) showing inversion of the mobility-death correlation suggested that contact rate might be dynamically regulated. To test this hypothesis, we developed a series of simplified Susceptible-Infective-Recovered-Deceased (SIRD) models to determine if instantaneous changes in contact rate and/or a dynamically changing contact rate (e.g., behavioral autorepression) could account for the observed inversion of the mobility-death correlation. Specifically, we modeled reductions in contact rates between individuals due to residential sequestration.

All SIRD models we utilize are based upon the same basic set of equations:

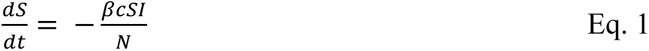

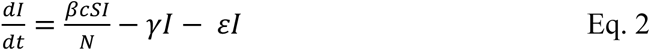

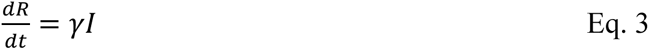

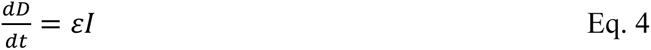

where S, I, R, and D represent susceptible, infectious, recovered, and deceased individuals in a population of N total individuals; *β* is the transmission rate constant (days^−1^), *γ* is the removal rate of infective individuals (days^−1^), *c* is the effective contact rate used to calculate “contact-reduction” in our models, and *ε* is the proportion of cases that result in death. The Susceptible-Infective-Recovered-Deceased (SIRD) model structure we chose was based on previous literature which expresses the death rate as a proportion of the number of infected individuals (27, 28). The different models we considered (below) differ only in the functional form of their contact rate.

The simplest SIRD model we considered (**Fig. 2A**) describes an extrinsically induced sequestration of individuals (e.g., government-mandated ‘lockdowns’) where the *c* is:

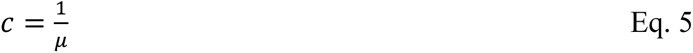

and *μ* is a constant that represents the efficacy of external forces (i.e., mandates) in facilitating the sequestration of healthy individuals (unitless). This initial model describes arguably the simplest constant reduction in contact rate (**Fig. 2B**). In this model, both initial and maximum reductions in contact rate reduced the number of calculated deaths (**Fig. 2C,D**), i.e., were negatively correlated with mortality, and could not reproduce the empirically observed inversion (**Fig. 1**).

**Fig. 2:**
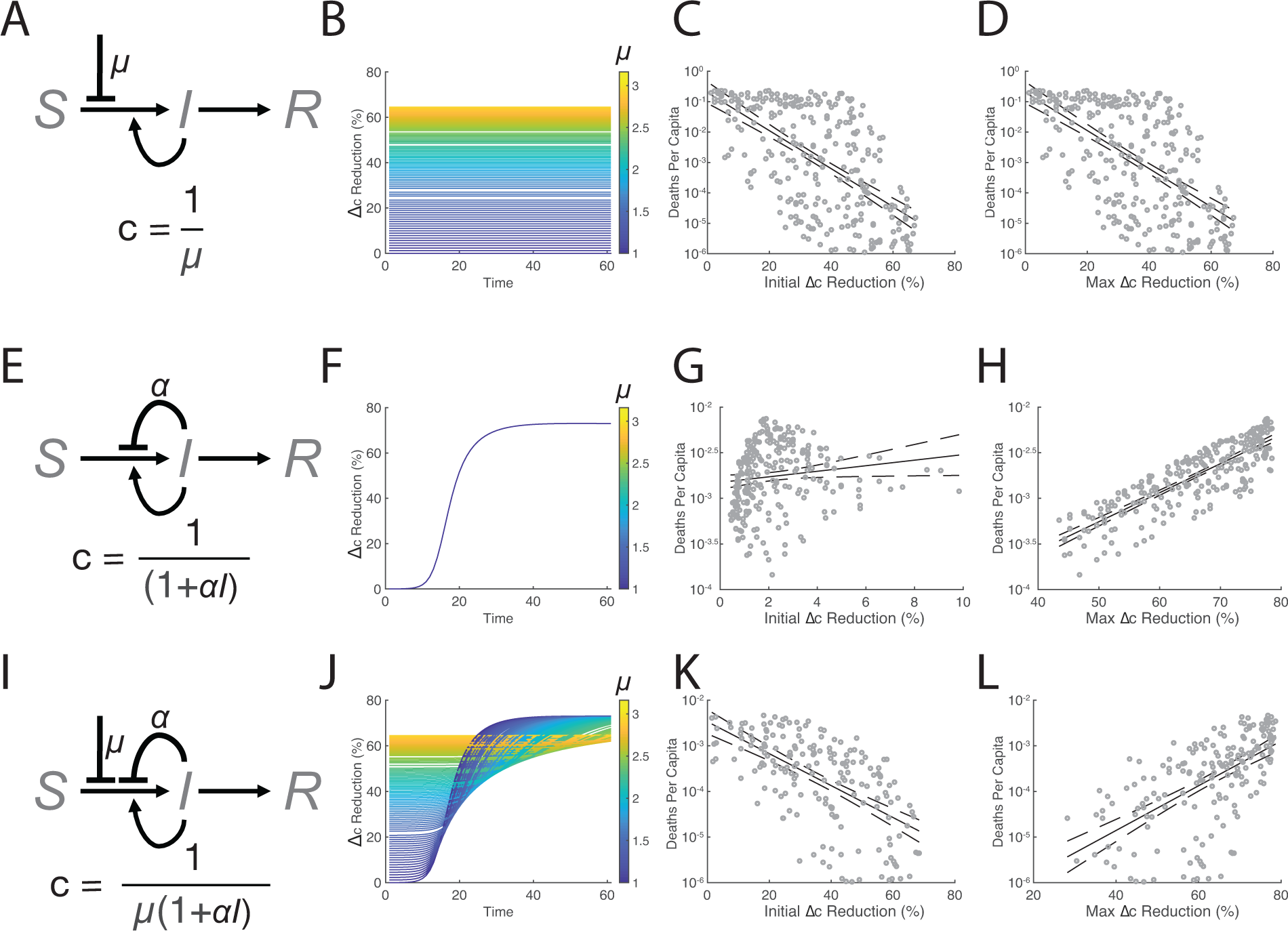
Autorepression coupled with sequestration is sufficient to account for temporal inversion of mobility-death correlation. **(A)** Schematic of the simple SIRD model [Eqs. 1-4] with extrinsic (e.g., policy based) sequestration (*μ*) [Eq. 5] where contact-reduction rate c = 1/*μ*. (**B**) Change in contact rate over time for simple SIRD (shown as % reduction). (**C-D**) Numerical simulations of mortality (calculated as 1% of R/N) as a function of initial and max changes in contact rate (% reduction) from simple SIRD model (regression p-values=1.6×10^−32^ and 1.6×10^−32^). (**E**) SIRD model with autorepression (*α*) [Eq. 6] where contact-reduction rate is c = 1/(1+*αI*). (**F**) Change in contact rate over time (% reduction) for SIRD autorepression model. (**G-H**) Mortality as a function of initial and max changes in contact rate (% reduction) from SIRD autorepression model (p-values=3.7×10^−2^ & 9.6 × 10^−69^, respectively). **(I)** SIRD model with coupled sequestration and autorepression [Eq. 7] where contact-reduction rate c = 1/*μ*(1+*αI*) (**J**). Change in contact rate over time for coupled SIRD extrinsic sequestration-autorepression model. (**K-L**) Mortality as a function of initial and max changes in contact rate (% reduction) for SIRD sequestration-autorepression model (regression p-values = 3.8×10^−20^ and 7.0×10^−20^, respectively). P-values indicate significance that the slope of the regression line is non-zero (null hypothesis slope=0).

Since this simple model [**Eqs. 1-5**] uses a fixed change in contact rate, to more closely mimic real-world scenarios of increases in strength of public-health mandates over time (29, 30), we also tested a variant of this model where induced sequestration was delayed and implemented at a time τ partway through the epidemic (i.e., a step function where *c*=1 if time<τ and c=1/μ if time≥τ). However, systems where induced sequestration was delayed still produced contact rate reductions that were negatively correlated with death (**Fig. S11**). Thus, a reduction in contact rate can only explain part of the correlation data (i.e., initial changes in residential occupancy and COVID-19 deaths), but not the temporal inversion of the correlation.

The second SIRD model (**Fig. 2E)** implements behavioral autorepression (i.e., negative feedback) by modifying the contact rate such that it reduces proportionally to the number of infectious cases:

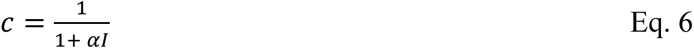

with *⍺* (person^−1^) representing the strength of the contact-rate reduction based on the number of infective individuals (a.k.a., the negative-feedback ‘gain’).

In this autorepression model [**Eqs. 1-4,6**], reductions in contact rate are solely driven by behavioral autorepression, such that residential occupancy increases in response to the number of infective individuals (**Fig. 2F**). The autorepression model generated a weak positive correlation between death and initial contact-rate reductions (**Fig. 2G**), but a strong positive correlation between deaths and maximum contact rate reductions (**Fig. 2H**). Thus, behavioral autorepression can also only explain a portion of the data—the correlation between deaths and maximum changes in residential occupancy—but not the negative correlation between death and initial increases in residential occupancy (**Fig. 2G,H**).

Based on these two sets of results each explaining a portion of the data, we constructed a third SIRD model combining both extrinsic sequestration and behavioral autorepression (**Fig. 2I**) with contact rate:

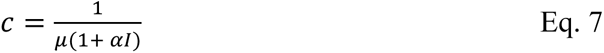

with all state variables and parameters as described above.

This combinatorial model [**Eqs. 1-4,7**] showed that regions with the weakest extrinsic (induced) sequestration *μ* (i.e., the least initial change in contact rate) generated the most infective individuals (as expected), but this ultimately resulted a larger contact-rate reduction later in time (**Fig. 2J**). Conversely, regions with the strongest extrinsic sequestration *μ* (i.e., the highest initial contact-rate reduction) generated the fewest infections, resulting in the lowest peak contact-rate reduction (**Fig. 2J**). This dynamic change in the effective contact rate was sufficient to generate a temporal inversion of the correlation between calculated deaths and contact-rate reduction (**Fig. 2K–L**), where mortality is negatively correlated with early contact rate reductions (**Fig. 2K**), but positively correlated with maximum contact-rate reductions (**Fig. 2L**). Overall, the simulations show that behavioral autorepression coupled with extrinsic (induced) sequestration is sufficient to recapitulate the inversion of the mobility-death correlation.

To test if a coincidental increase between residential occupancy and COVID-19 cases could have caused the inversion of the mobility-death correlation, we simulated a scenario where changes in residential occupancy did not reduce contact rates, but found this scenario could not cause the temporal inversion (Supp Text-Section 2), arguing that a functional dependency, not a coincidental increase, between contact rate and infection is required (**Fig. S12**).

To test alternate hypotheses that could account for the inversion of the mobility-death correlation, we considered the possibilities that intense lockdowns drove infection, or that deaths (rather than infections) drove changes in behavior (Supp Text-Section 3). However, neither alternate hypothesis was parsimonious with the epidemiological data (**Fig. S13–S14**).

### Fluctuation analysis reveals signature and strength of autorepression

To verify the existence of negative feedback in these early COVID-19 epidemiological dynamics and quantify the feedback strength, we exploited a known phenomenon where negative feedback typically reduces the magnitude of fluctuations in a system, proportional to the feedback strength (31–33). To search for this effect, we analyzed variance in daily COVID-19 infection data over time using a common metric; the normalized variance (i.e., square of the standard deviation divided by the mean, also known as the ‘Fano factor’) of daily infections.

First, a stochastic SIR model (**Table S1**) was used to benchmark the analysis, with Monte-Carlo simulations (34) used to examine how behavioral autorepression (**Fig. 3A**) affected the normalized variance of the daily infection rate in the model. As expected, models incorporating autorepression showed a marked reduction in the Fano factor versus mean infections per day (**Fig. 3B**). Specifically, the analysis indicated that incorporating autorepression in an SIR model causes the slope of the Fano to decrease.

**Figure 3:**
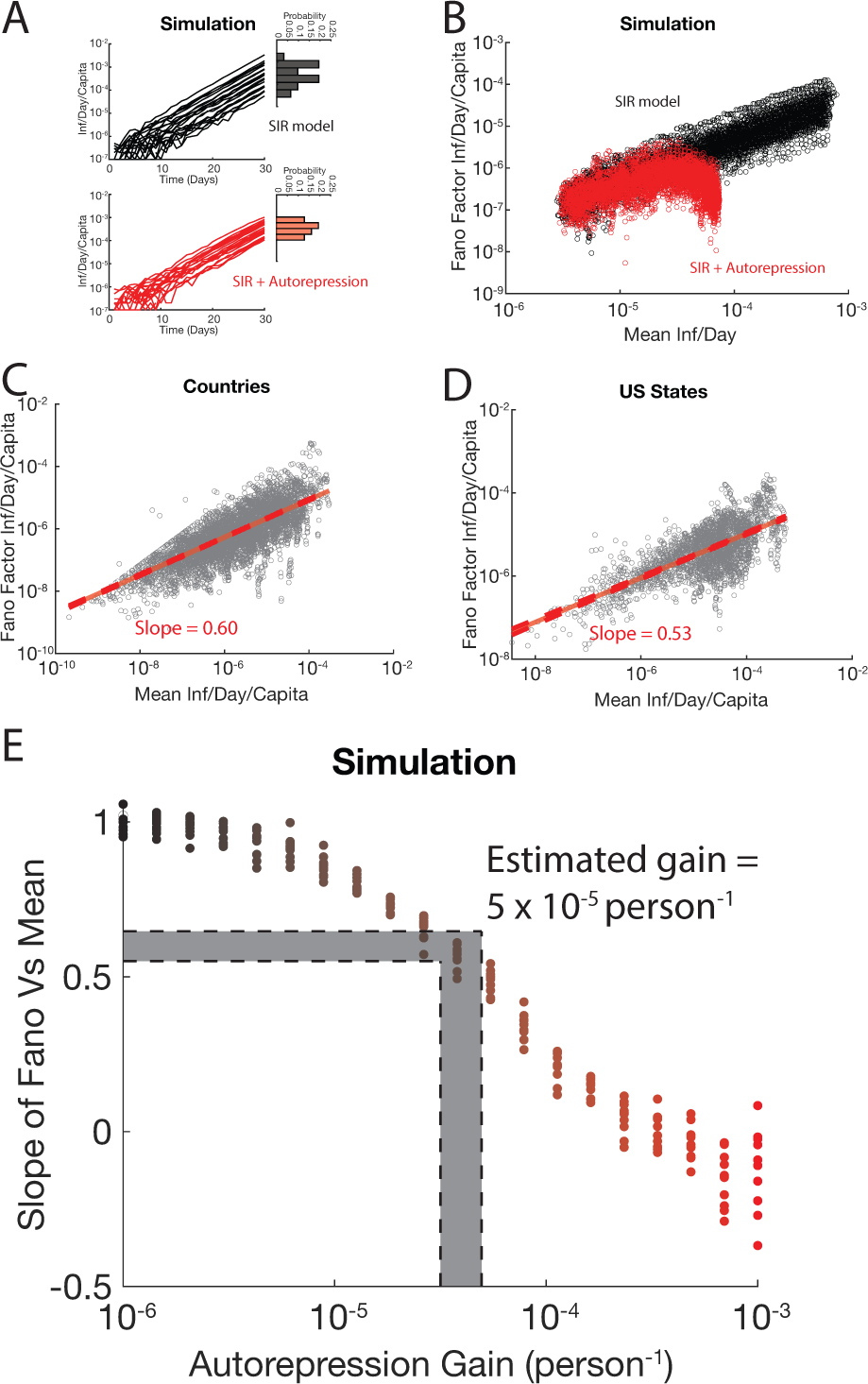
Variability in infection rate can be used to verify autorepression and calculate its strength. (**A**) Representative Monte-Carlo simulations of infection rate (infections per day) over time in a stochastic SIR model without autorepression (black) and with autorepression (red). Histograms show the variability in infections per day at day 30. (**B**) Fano factor (variance normalized to mean) of infections rate versus mean generated by Monte-Carlo simulations of an SIR model with (red) and without (black) autorepression. (**C**) Data of Fano factor versus mean of the first 60 days of infection for countries affected by COVID-19. Slope of linear regression = 0.6 (dashed red lines represent 95% confidence interval). (**D**) Fano factor versus mean plot of the first 60 days of infection for US states affected by COVID-19. Linear regression slope = 0.53 (dashed red lines represent 95% confidence interval). (**E**) Effect of autorepression strength on slope of the Fano factor vs mean. The slopes calculated from state and country data correspond to an autorepression gain of approximately 5 × 10^−5^ person^−1^. Shaded area represents mean vs. Fano slope and corresponding autorepression gain range determined by measurements at the international and US state scale.

Next, we quantified the reduction in the empirical Fano of the COVID-19 infection data by least-squares linear regression for daily infections early in the epidemic internationally (**Fig. 3C**) and in the United States (**Fig. 3D**). The slope of the Fano-vs-mean relationship in these datasets was found to be ∼0.6, consistent with autorepression (**Fig. 3C,D**).

To quantify the negative-feedback strength (a.k.a., negative ‘gain’) of behavioral autorepression, we generated a series of simulated slopes from models with incrementally stronger autorepression (**Fig. 3E**). As the simulated strength of autorepression increased, the Fano-versus-mean slope decreased, and extrapolation to the fit US state and country data estimated the autorepression gain for the COVID-19 pandemic at 5×10^−5^ person^−1^ (**Fig. 3E**). An autorepression gain of 5×10^−5^ person^−1^ indicates that when the level of infection reaches two thousand simultaneously infective individuals, the transmission rate constant in that outbreak will be decreased by 10%.

While the decreasing Fano-versus-mean slope could in principle be accounted for by mechanisms other than autorepression (e.g., policy changes that reduce transmission rate), the modeling results above (**Fig. 2, Fig S.11**) suggest that a feedback mechanism is the most parsimonious with the data. Moreover, analysis of early changes in policy (35) were found to be positively correlated with infection count (**Fig. S15**), suggesting policy itself may play a role in behavioral autorepression.

To test if pre-emptive, policy-mediated modification of the transmission rate constant could account for infection-rate variability, we analyzed simulations where the transmission rate constant was varied (**Fig. S16**). Changing the transmission rate constant could increase the Fano-vs-mean slope above 1, but could not reduce the Fano-vs-mean slope below 1, again supporting the finding that a reduction in the Fano slope is a signature of autorepression (**Fig. 3, Fig. S16**). Notably, quantification of the Fano-vs-mean relationship over the first 15 days of infection at the international (**Fig. S17A**) and US state (**Fig. S17B**) scale also yielded slopes below 1. Since COVID-19 mortality occurs ∼20 days after exposure to SARS-CoV-2 (36), these results suggest a substantial fraction of variation in infection counts is suppressed by infection-induced autorepression independently of death-induced autorepression.

### Temporal analysis of infection waves indicates presence of autorepression

To establish an independent line of evidence for behavioral autorepression, we qualitatively analyzed the dynamics and temporal characteristics of the initial ‘wave’ of COVID-19 infection in different regions. Specifically, we analyzed if the wave pattern (i.e., magnitude and timing of infection waves) was consistent with a traditional SIR model [**Eqs. 1–4**] or whether incorporating delayed autorepression [**Eq. S2**] was more parsimonious with the data (**Fig. S18A,** Supp Text-Section 4).

The SIR model [**Eqs. 1–4**] naturally produces an initial wave, or peak in infections per day, and we first benchmarked how modulating the infection rate (*β*) affected this wave (**Fig. S18B**). As expected, increased infectivity generated earlier and larger peaks in infections per day (**Fig. S18C**), producing a trend where earlier waves peaked at higher levels and later waves peaked at lower levels. Similarly, when a fixed time delay (*τ*) was incorporated between the susceptible and infective populations (**Fig. S18D, Eqs. S3-S6**), increasing this delay generated a similar trend where infection waves peaked later and at lower peak magnitudes (**Fig. S18E**).

In contrast, when behavioral autorepression (**Fig. S18F, Eq. S2**) was incorporated, a starkly different trend emerged: increasing the delay time generated stronger waves that peaked later but at *higher* magnitudes (**Fig. S18G**). This observation held true for a range of different parameter values.

To compare these model trends to the empirical data, we calculated timing of COVID-19 infection waves (i.e., days from a region’s epidemic onset to its first peak in infections/day), and compared it to the magnitude of that region’s first wave. The data at both the international and US-state scales showed an increasing trend (i.e., positive correlation) between the magnitude and the timing of the peak with larger peaks occurring at later times (**Fig. S18H & S18I**; p-value: 2×10^− 3^ & 2×10^−9^ respectively). Overall, these data appear qualitatively consistent with autorepression and suggest that temporal delays in autorepression vary across different locations.

### Impacts of behavioral autorepression on epidemic forecasting

To determine the impact of behavioral autorepression on epidemic forecasting, we mirrored previous forecasting approaches (37), using the first 60 days of reported US COVID-19 deaths to predict the subsequent outbreak trajectory using either a canonical SIRD model [**Eq. 8-11**] or a behavioral autorepression SIRD model with autorepression gain determined from the first 60 days of data [**Eq. 12-15**]. The equation sets used are analogs of Eqs. 1–4 (with Eq. 5 and 7 respectively), and are as follows, for the canonical SIRD model:

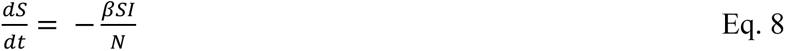

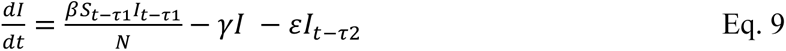

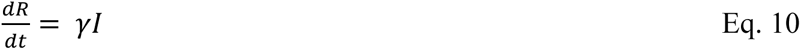

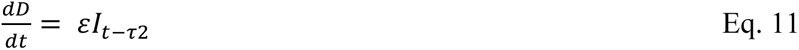

and, as follows, for the autorepression model:

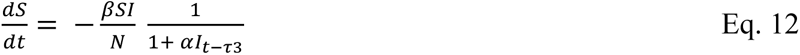

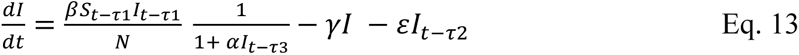

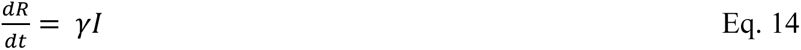

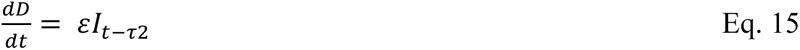

with *τ*_1_(days) representing the time it takes for a susceptible individual exposed to SARS-CoV-2 to become infectious, with *τ*_2_(days) representing the delay between an individual becoming infectious and that individual dying from COVID-19, and *τ*_3_(days) representing the time it takes an increase in COVID-19 cases to cause susceptible individuals to self-sequester and mediate autorepression. All other state variables and parameters as described above.

Each model (either **Eqs. 8–11** or **Eqs. 12–15**) was “trained” on the first 60 days of data on COVID-19 deaths in the US using nonlinear least squares regression analysis and simulations, and then extended out to 120 days for prediction. As previously reported (37), the SIRD model [**Eqs. 8–11**] forecasted ∼10^6^ COVID-19 deaths within the first 120 days of the epidemic, whereas the reported COVID-19 deaths were substantially lower in this time period. In contrast, the autorepression model [**Eqs. 12–15**] generated forecasts of deaths over 120 days that appeared far more parsimonious fits with the data than the simple SIRD model forecast (**Fig. 4A**). To quantitatively compare the goodness of fit between the two models, the Akaike Information Criterion (AIC) metric was used (38), and the SIRD forecast had a significantly poorer score than the autorepression model (AIC = 145 vs –322; Wilcox rank sum test, p-value: 0). Since lower AIC represents a better fit, these results demonstrate that inclusion of behavioral autorepression in epidemiological model formulation significantly improves the forecasting potential of these models.

**Figure 4:**
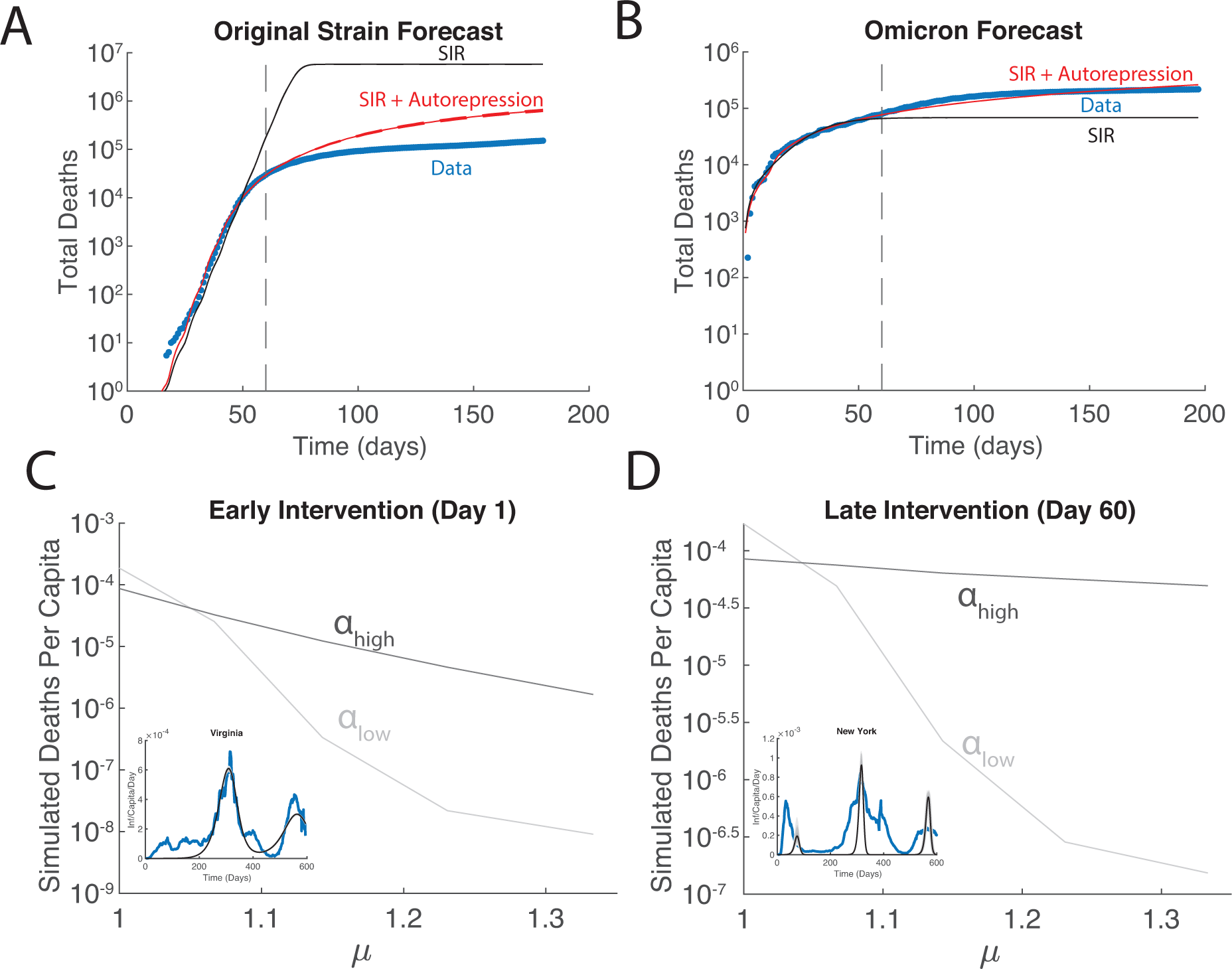
Autorepression models significantly improve COVID-19 forecasting. **(A)** Mortality forecasts of COVID-19 deaths in first 180 days after the COVID-19 pandemic initiation in the US. COVID-19 morality data (blue), forecasted death from the standard SIRD model (black) and forecasted death from SIRD-autorepression model (red) showing that autorepression model forecasts are more accurate than canonical SIRD model forecasts (SIR AIC=145 vs. AIC=–322 for SIR-autorepression) **(B)** Mortality forecasts of COVID-19 deaths after Omicron variant outbreak (first 200 days) in the US. COVID-19 morality data (blue), forecasted death from the standard SIR model (black) and forecasted death from SIRD-autorepression model (red) showing that autorepression model forecasts are more accurate than canonical SIR model forecasts (SIRD AIC=–398 vs. AIC=–978 for SIR-autorepression) **(C)** Forecasted efficacy of non-pharmaceutical intervention (lockdowns) when initiated early in the epidemic for varying changes in extrinsic (e.g., policy based) sequestration (*μ*) under scenarios of low or high autorepression strength (alpha). Mortality totals were simulated based on fits to COVID-19 infection dynamics. Intervention strength based on mobility changes from Fig. 1. Light grey-weak fitted autorepression gain, dark grey-strong fitted autorepression gain. Insets show examples of fitting, blue represents smoothed infections per day, black line represents averaged dynamics from autorepression model, grey shading represents 99% confidence interval. **(D)** Forecasted efficiency of non-pharmaceutical intervention initiated late in epidemic (day 60) under scenarios of low-vs.-high autorepression strength (alpha).

The forecasting potential of the behavioral autorepression SIRD model was further verified using the first sixty days of the omicron wave. Autorepression models forecasted total mortality with a high degree of accuracy over ∼140 days, while SIRD models generated forecasts that severely underpredicted mortality, displaying a significantly poorer AIC score (**Fig. 4B** AIC = - 398 vs -978, Wilcox rank sum test, p-value: 0). These results further support the utility of epidemiological models incorporating behavioral autorepression, and suggest that these models could be used to make health policy decisions early on during a pandemic.

To determine the impact of behavioral autorepression on the efficacy of non-pharmaceutical interventions (NPIs), a series of NPIs were simulated. To obtain realistic parameters for these simulations, autorepression models were fitted to infections per day from all 50 states. These fitted parameters were then used to run data-driven simulations of NPIs that reduced the SARS-CoV-2 transmission rate constant by 0–25% (i.e., the range of mobility changes empirically measured during the first wave of the pandemic, see Fig 1). As the NPI efficacy metric, we used number of deaths averted. The simulations showed that while two regions may have similar mortality totals, the effectiveness of intervention differs widely depending on the autorepression gain of an individual region (*⍺* between 4.1 × 10^−6^ and .32 person^−1^). Simulations of regions where autorepression is weak (example fit in **Fig. 4C** inset) had dramatic responses to early NPIs, represented in the graph by simulations that show an ∼4 log-fold decrease in deaths in response to a NPI that decreases transmission by 25% (**Fig. 4C** light grey line). Over the same range of NPI strengths, simulations of regions where autorepression is strong had ∼1 log-fold reduction in death (**Fig. 4C** dark grey line). Late implementations of the same NPIs yielded an ∼3 log-fold decrease in deaths in regions when autorepression is weak and < 0.5 log-fold decrease in regions when autorepression is strong (**Fig. 4D**, example fit in **Fig. 4D** inset). These results predict that NPIs are less effective in regions with high autorepression gains. Suppression of variation is a well-known phenomenon in autorepression systems, caused by their capacity to self-regulate (31, 32, 39). These results suggest that autorepression models could be used to forecast the efficacy of NPIs in a region-specific manner.

## DISCUSSION

This study presents multiple pieces of evidence supporting the presence of negative-feedback dynamics (i.e., behavioral autorepression) during the early stages of the COVID-19 pandemic. Simulations using minimal models demonstrated that behavioral autorepression is sufficient to explain a statistically significant inversion of the correlation between residential occupancy and COVID-19 mortality over time that was consistently observed across epidemiological scales (**Figs. 1–2**). Second, fluctuation analyses of SARS-CoV-2 daily infection data was consistent with autorepression’s ability to reduce the magnitude of fluctuations and enabled estimation of the negative-feedback strength for the COVID-19 pandemic at 5×10^−5^ person^−1^ (**Fig. 3**). This value indicates that when the level of infection reaches two thousand simultaneously infective individuals, the transmission rate constant in that outbreak will be decreased by 10%. Importantly, the majority of the reduction in fluctuations was observed during very early times (i.e., <= 15 days; **Fig. S16**), indicating that while death-mediated autorepression (27) may exist, infection-mediated autorepression had an independent, substantial impact on SARS-CoV-2 infection dynamics. Third, temporal analysis of the first wave of infection revealed that variation between peak timings and intensities is parsimonious with autorepression delay rather than regional variation in infectivity (**Fig. S17**). Taken together, these analyses provide independent lines of evidence for the impact of behavioral autorepression on SARS-CoV-2 transmission and indicate that the inclusion of autorepression in models can significantly improve epidemiological forecasting and may affect the efficiency of certain NPIs (**Fig. 4**).

Interestingly, the behavioral autorepression model indicates that mobility, in particular residential occupancy, is unlikely to be an independent variable, at least early during an outbreak. Rather, the models indicate that mobility measurements, a surrogate of contact rate, are in fact an ensemble variable influenced by extrinsic factors (e.g., public-health mandates) as well as dynamic individual factors (e.g., risk-taking). Several studies have reported on the efficacy of early lockdowns (40–42), corroborating the observation that initial changes in residential occupancy are negatively correlated with regional mortality. The results presented here also help account for the previously-unexplained observation that early lockdowns were more effective than late lockdowns (41, 42), since the models argue that mobility changes induced by late lockdowns become overwhelmed by collective decisions to self-sequester in response to infection reports.

Perhaps the most utilitarian impacts of the presence of behavioral autorepression are its effects on the fidelity of epidemic forecasting and the efficiency of NPIs. The COVID-19 pandemic highlighted substantial variation in epidemiological forecasts (37, 43), likely due to numerous factors including unknown parameter estimates, assumptions about efficacy of NPIs, and model structure. This study demonstrates that SIR models incorporating behavioral autorepression can generate less variable forecasts, and that the behavioral autorepression phenomenon may aid in judicious application of NPIs. The finding that autorepression affects NPI efficacy suggests that measuring local variation in autorepression strength could allow NPIs to be tailored to individual regions, assuming that autorepression strength is intrinsic to a population and does not substantially change over time or between outbreaks, which will require further study (see discussion below). Since autorepression is often correlated with oscillation (9, 13, 14, 27), future models could, in principle, accurately forecast the timing of waves of COVID-19 infection.

Future work concerning behavioral autorepression during outbreaks of infectious disease should address the consistency of autorepression parameters within outbreaks and between different outbreaks. Our study treated autorepression as constant during the course of the COVID-19 pandemic to identify the minimum conditions sufficient to explain the changes in correlation, variability, and timing observed in infection trajectories. However, it is possible that as public understanding of COVID-19 evolved, autorepression strength changed. This work similarly does not investigate whether previous pandemics experienced similar levels of autorepression. However, since behavioral autorepression was originally hypothesized to explain saturation of cholera infection rate (2) rather than SARS-CoV-2, it seems possible that autorepression is an intrinsic property of human populations experiencing an outbreak rather than a property of the pathogen driving an outbreak. Therefore, analysis of autorepression during the COVID-19 pandemic could serve as a guide to studying autorepression and its consequences in future pandemics.

## METHODS

### Linear regression analysis

Linear regressions were performed in MATLAB using the fitlm function. Linear regressions were performed using changes in residential occupancy to explain regional variation in confirmed COVID-19 deaths (20). Changes in residential occupancy were measured using the Google COVID-19 Community Mobility Reports (21). COVID-19 death totals on 05/24/2020 were compared to the change in occupancy from baseline after a threshold of ten deaths had been crossed (termed “*initial change in residential occupancy*”) and the maximum change in residential occupancy during the first wave of COVID-19 infection (termed “*maximum change in residential occupancy*”). 05/24/2020 was chosen as the cutoff date for quantifying COVID-19 deaths, in part, in order to avoid confounding deaths during the first wave of infection with deaths from subsequent waves. COVID-19 death totals were Log transformed prior to regression and regression analysis performed at three different geographic scales based on available data, including: international countries; US states, and US counties. Unless specifically noted, linear models only have one independent variable and one dependent variable. Multi-factor linear models have their individual independent variables listed in their corresponding figure legends.

### ODE simulations of policy- vs. feedback-controlled epidemics

Nonlinear ordinary differential equations were numerically solved in MATLAB using ode15s. Initial parameters were randomly generated from empirically chosen ranges that produced >1 infection but infected <50% of the population in sixty days. Two hundred distinct parameter sets were simulated to represent a set of different regions with unique epidemiological parameters. Percentage reduction from initial contact rates to contact rates at time t were calculated via **Eq. 16**:

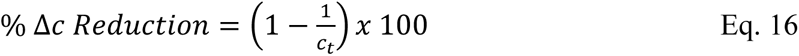

where *c_t_* corresponds to the contact rate at time t. Each simulated region was analyzed to find the early change in contact rates and the maximum change in contact rates. The early change in contact rates (*c_e_*) was calculated via **Eq. 17**:

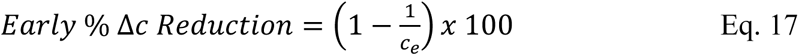

where *c_e_* corresponds to the contact rate when a threshold of ten deaths was reached. The maximum change in contact rates (*c_m_*), was calculated using **Eq. 18**:

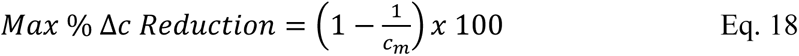

where *c_m_* corresponds to the maximum change in contact rate over the course of the simulation. These percentage changes in contact rates were used to calculate a linear regression between Δ*c* and simulated deaths using the same procedure as Methods-Linear regression analysis. When policy was implemented partway through the simulation, it was treated as a step change in the mandate variable *μ* that occurred after thirty days of simulation.

### Monte Carlo simulation of infections per day

A Monte Carlo algorithm for MATLAB was used (https://github.com/nvictus/Gillespie) to implement a stochastic SIR model. Simulations were run over a sixty-day time period. Simulation parameters were empirically determined to match the first sixty days of infection dynamics from countries and US states. The variance and mean of individual infection-per-day trajectories were measured along seven-day moving time windows. In each window, the variance was divided by the mean to calculate the Fano factor (i.e., variance normalized by mean) for infections per day.

### Perturbation analysis of SIRD versus autorepression models

Randomized parameter sets were generated from an empirically-defined range chosen to maximize simulated epidemics that produced >1 infection but infected <50% of the population in sixty days. Epidemics were simulated for 3,650 days (∼10 years) to ensure that infections per day reached a peak during the simulation time. Simulated epidemics that failed to generate more >1 infection or infected >50% of the population in sixty days were excluded from the analysis, whereas simulations that generated >1 infection and infected <50% of the population in sixty days were marked for analysis. Simulated epidemics marked for analysis were analyzed for their wave dynamics by using the findpeaks function in MATLAB. The time until the first peak in infections per day and the number of infections on that day were quantified.

### Fitting and forecasting COVID-19 mortality dynamics

MCMC fitting was performed in MATLAB (https://github.com/mjlaine/mcmcstat) using confirmed COVID-19 deaths in the United States for first 60 days after the first US COVID-19 infection was recorded. Hand fitting was used to initialize parameters before MCMC fitting. Sum of squared error was used to optimize the fit within the algorithm. Four independent fits were performed, with 10,000 iterations per fit. The parameter set from best fit out of the four independent runs over the initial 60-day time period was used to forecast mortality totals over the next 120 days (a total simulation time of 180 days). Confidence interval projections and Akaike Information Criterion (AIC) scores were calculated using the ODE values generated by parameter sets from the final 1000 iterations in the MCMC fitting process. AIC scores were calculated using **Eq. 19**:

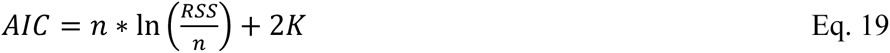

where *n* is the number of data points used in the fitting, RSS is the residual sum of squares, and *K* is the number of fitted parameters.

Individual AIC scores were generated for each of the final 1000 iterations, then averaged to represent the AIC score of a fit.

### Fitting daily COVID-19 infection dynamics

MCMC fitting was performed in MATLAB (https://github.com/mjlaine/mcmcstat) using daily counts of confirmed COVID-19 infections at the international or US state level from the first day an infection was recorded in a given region until October 26^th^, 2021. Hand fitting was used to initialize parameters before MCMC fitting. **Eqs. 1-4** and **Eq. S2** were used to generated simulated infections per day, which was compared to real infections per day. Sum of squared error was used to optimize the fit within the algorithm. 60 independent fits were performed per state with 10,000 iterations per fit. The best fit was chosen by the average AIC (calculated using **Eq. 19**) of the final 1000 iterations for each state.

### Forecasting intervention efficacy

Epidemic dynamics were simulated using a modified Euler method for SDE simulation where infective individuals exerted delayed autorepression on epidemic growth. Parameters were taken from the converged fits of the delayed behavioral autorepression model for infection dynamics in all fifty US states. Simulations represented one year of mortality dynamics. Interventions that statically decreased the transmission rate constant (*β*) were applied at day 1 or day 60 to represent early and late interventions respectively.

## Data Availability

All data produced in the present study are available upon reasonable request to the authors

## Abbreviations

AIC: Akaike Information Criterion
COVID-19: COronaVIrus Disease of 2019
SIR: Susceptible-Infective-Removed
SIRD: Susceptible-Infective-Recovered-Deceased
NPI: Non-Pharmaceutical Intervention

## AKCNOWLEDGEMENTS

We thank Katie Claiborn for editing, Rob Rodick for discussions on the effect of population density on the spread of COVID-19, and Brad Pollock for guidance on statistical methods. This work was supported by NIH grant R37AI109593.

## SUPPLEMENTARY TEXT

### Section 1 Verifying robustness of mobility-death inversion

To determine if potential covariation between the early and maximum mobility metric could account for the inversion of the mobility-death correlation, we used linear regression models that account for the simultaneous effect of population, initial residential occupancy, and maximum residential occupancy on death (44). These multi-factor regressions recorded the same negative correlation between death and initial residential occupancy and the same positive correlation between death and maximum residential occupancy (**Fig. S7A-B**). Thus, the inversion of the mobility-death correlation does not appear to be dependent on covariance between population and mobility or covariance between mobility metrics (**Fig. S7A-C**).

Since regional population size is known to be correlated with density (45) and SARS-CoV-2 transmission is known to be density-dependent (46), we next asked if a (nonlinear) quadratic relationship between population size and death rates could account for the inversion of the mobility-death correlation. Models assuming a quadratic dependence between population size and death also fit well (**Fig. S8A**), but the death residuals still exhibited an inversion of the correlation between initial and maximum residential occupancy (**Fig. S8B-C**). Thus, models that account for the nonlinear relationship between population size and deaths further support the inversion of the mobility-death correlation.

To ensure that the inversion of the mobility-death correlation was not unique to Google mobility data, we repeated the analyses using an alternate population mobility dataset (47) that used aggregated navigation data from Apple Maps. Single-factor linear regressions between mobility and death showed the expected correlation between initial change in mobility and death (**Fig. S9A**), but did not show the counterintuitive correlation between the maximum change in mobility and death (**Fig. S9B**). A multifactor linear regression that accounts for covariation between population, initial change in mobility, maximum change in mobility, and death showed that the correlation between death and mobility inverts over time (**Fig. S9C-E**). These results demonstrate that inversion of the mobility-death correlation is not unique to the Google mobility dataset (**Fig. S9**).

To confirm that the counterintuitive correlation between maximum change in residential occupancy and death was robust and not due to a specific instantaneous measure of maximum mobility, we analyzed cumulative change in mobility. The cumulative changes for most mobility measures (residential, retail, grocery, transit, and workplace occupancy) showed similar positive correlation with deaths per capita at all geographic scales (**Fig. S10**), although park occupancy showed no significant correlation with deaths per capita. Overall, cumulative changes in mobility appear to exhibit the same correlation with deaths per capita as the maximum instantaneous change in mobility.

### Section 2 Coincidental rise in residential occupancy and infections does not cause inversion of the mobility-death correlation

This SIR model simulates outbreaks without repression, representing a scenario where residential occupancy values are not affected by induced sequestration or autorepression and do not decrease contact rates amongst the population (**Fig. S12A, Eq. 1-4, Eq. S1**). Residential occupancy values were randomly drawn from a set of increasing contact rate reduction values (**Fig. S12B**) and compared against epidemics that were simulated without any reduction in transmission. Neither early nor maximum changes in residential occupancy were correlated with death (**Fig. S12C,D**). These results suggest that coincidental increases in residential occupancy during the onset of the COVID-19 pandemic are not sufficient to generate the correlations observed in the analyses performed in this study (**Fig. 1**).

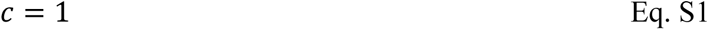

*c* represents the effect of contact rate on transmission based on sequestration of healthy individuals (unitless).

### Section 3 Investigating hypotheses to explain mobility-death inversion

The correlation between initial changes in residential occupancy and reductions in COVID-19 mortality (**Fig. 1** 1^st^ column) is consistent with previous research showing that fast lockdowns reduce disease transmission (10, 40, 42). These studies are consistent with the observation that contact frequencies go down under conditions where residential occupancy increases (26). However, these studies cannot explain the positive correlation observed between maximum changes in mobility and COVID-19 deaths (**Fig. 1** 2^nd^ column). Therefore, we set out to test hypotheses that could potentially explain this positive correlation.

To test whether lockdown-mediated increases in household contacts led to enhanced transmission overall, we used US housing density data (proportion of households with > 1.5 occupants/room) to search for a synergistic effect between density and mobility on regional mortality. Previous works have shown that SARS-CoV-2 transmission is density-dependent (46), household transmission is a major component of COVID-19 infection (48), and that lockdown-specific changes in contact patterns can change transmission on a population scale (49). To assess the impact of household transmission during lockdown, we performed a partial least squares regression (44) accounting for individual contributions of density, population, initial change in mobility, maximum change in mobility, and a density-mobility interaction on COVID-19 mortality totals. The density-mobility interaction term was calculated by multiplying the proportion of households with > 1.5 occupants/room by the maximum change in residential occupancy. With this regression approach, we were able to deconvolve the individual effects of density and mobility from the interaction between the two factors. If transmission of COVID-19 had been increased through increased occupancy of dense households, the multi-factor regression should reveal a synergistic interaction between population density, maximum change in residential occupancy, and COVID-19 deaths. The analysis showed no significant interaction (p-value: 0.25) between housing density and maximum change in residential occupancy (**Fig. S13**), suggesting that lockdowns do not exacerbate overall transmission through increased household contacts. Notably, this lockdown-mediated transmission hypothesis is also difficult to reconcile with the expected correlation observed between initial changes in mobility and deaths per capita (**Fig. 1**).

To determine whether the counterintuitive correlation between reduced mobility and increased COVID-19 deaths could be caused by an inverted causation-correlation relationship (27) (i.e., that increased death was the driver of reduced local mobility), we examined whether changes in death were required to change mobility on a region-to-region basis. Analysis of changes in mobility prior to a single reported death (“pre-death” mobility changes) showed that residential occupancy typically increased ∼10–15% before a single death (**Fig. S14**). Since the maximum increase in residential occupancy for any region was ∼20-30%, these results show that countries, states, and counties were halfway to peak shutdown before a single local death occurred. This substantial change in residential occupancy before the onset of death suggests that while COVID-19 deaths may contribute to changes in population-level behavior, local deaths are unlikely to be the driving force behind increased residential occupancy. Moreover, this death-mediated-mobility hypothesis is difficult to reconcile with the expected correlation between initial changes in mobility and deaths per capita.

### Section 4 Delayed autorepression is used to analyze infection waves

Delayed autorepression was represented by a change in the contact rate, c, where:

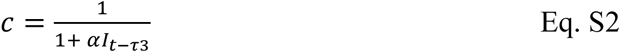

with *⍺* (person^−1^) representing the strength of the contact-rate reduction based on the number of infective individuals (a.k.a., the negative-feedback ‘gain’) and *τ*_3_ (days) representing autorepression delay.

Delayed infectivity was represented by the following set of equations:

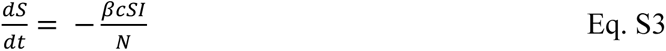

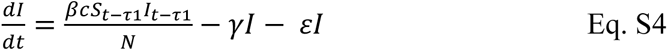

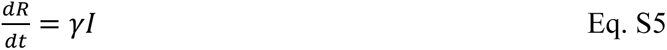

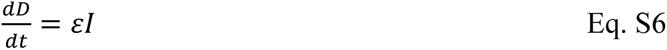

where S, I, R, and D represent susceptible, infectious, recovered, and deceased individuals in a population of N total individuals; *β* is the transmission rate constant (days^−1^), *γ* is the removal rate of infective individuals (days^−1^), *c* is the effective contact rate used to calculate “contact-reduction” in our models, and *ε* is the proportion of cases that result in death, *τ*_1_ (days) is the time it takes for a susceptible individual exposed to SARS-CoV-2 to become infectious.

**Table S1:** Reactions for a stochastic model of behavioral autorepression.

| Reaction | Rate of Reaction | Description |
| --- | --- | --- |
| $S \rightarrow I$ | $\frac{\beta S I}{1 + \alpha I}$ | Infection of a susceptible individual under autorepression control |
| $I \rightarrow R$ | $\gamma I$ | Removal of individual from infective population when individual recovers or becomes deceased |

### Supplementary Methods

#### Multi-factor regression of interaction between residential occupancy and housing density

Multi-factor linear regressions were performed using fitlm in MATLAB. These regressions were performed using housing density, population, initial changes in residential occupancy, and maximum changes in residential occupancy, and the interaction between maximum changes in residential occupancy and housing density to explain regional variation in confirmed COVID-19 deaths among US counties. Housing density was measured through US census reports on the proportion of households in a county that had more than 1.5 occupants per room (50). County populations were also measured using counts from the US census (50). Confirmed COVID-19 death totals on May 24^th^, 2020 were used to quantify pandemic mortality (20). The logarithm of COVID-19 death totals was taken before analysis.

Changes in residential occupancy were measured using the Google COVID-19 Community Mobility Reports (21). Early changes in occupancy from baseline were quantified by measuring the change in residential occupancy after a threshold of ten deaths had been crossed (termed “*initial change in residential occupancy*”). Later changes in occupancy from baseline were quantified by measuring the maximum change in residential occupancy during the first wave of COVID-19 infection (termed “*maximum change in residential occupancy*”). The product of the maximum change in residential occupancy and housing density on a county-by-county basis represented the interaction between increased indoor occupancy and density-based transmission. When the multifactor linear regression corrects for linear contributions of residential occupancy and housing density, the coefficient and significance of the interaction term represents the strength of a potential synergistic effect between increased indoor occupancy and density-based transmission.

---

**Supplementary Figure 1:**
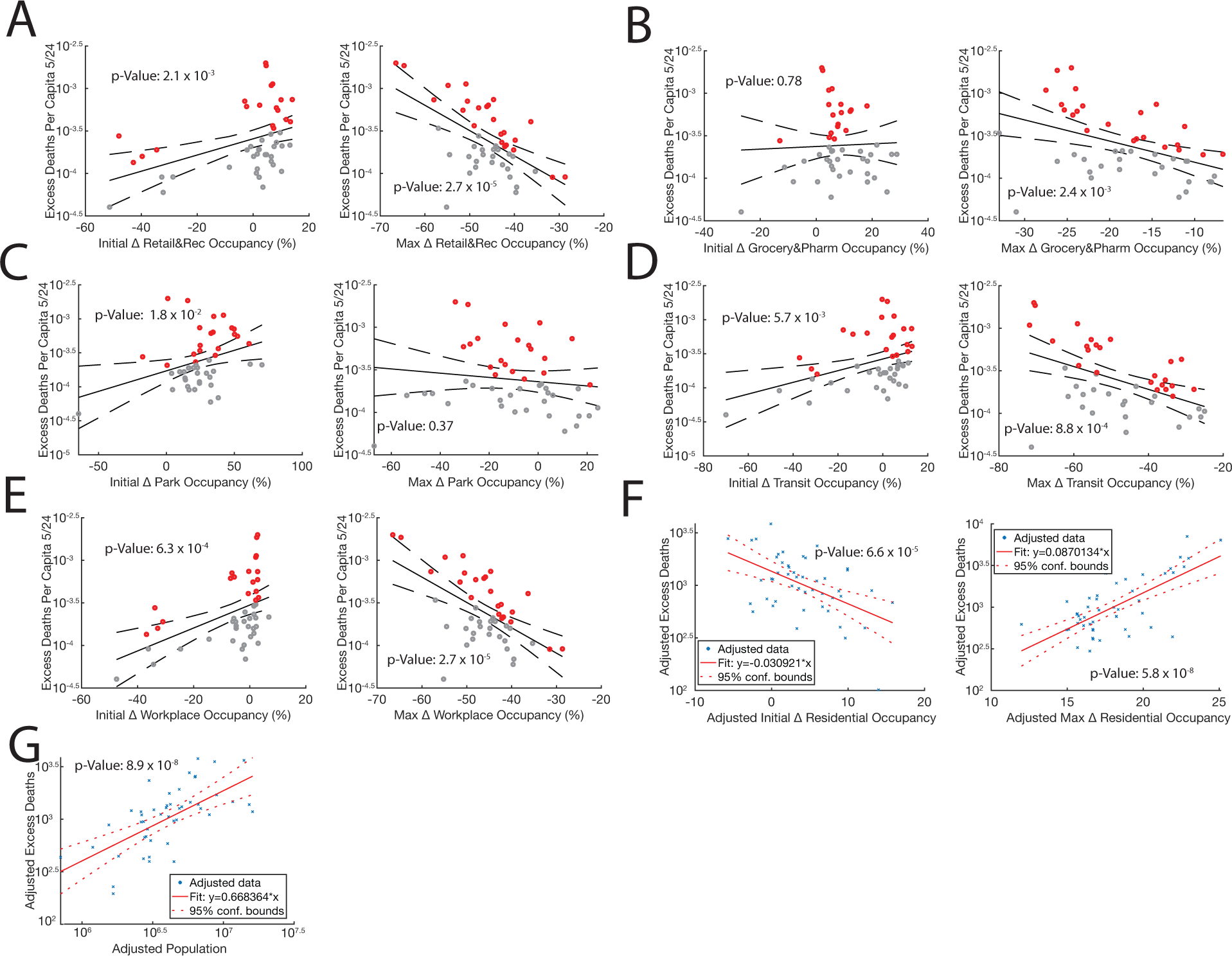
Occupancy and excess deaths exhibit correlation inversion. A. (1^st^ column) Linear regression of the initial change in retail and recreation occupancy versus the excess deaths per capita of all states affected by COVID-19. (2^nd^ column) Linear regression of the maximum change in retail and recreation occupancy versus the excess deaths per capita of all states affected by COVID-19. B. (1^st^ column) Linear regression of the initial change in grocery and pharmacy occupancy versus the excess deaths per capita of all states affected by COVID-19. (2^nd^ column) Linear regression of the maximum change in grocery and pharmacy occupancy versus the excess deaths per capita of all states affected by COVID-19. C. (1^st^ column) Linear regression of the initial change in park occupancy versus the excess deaths per capita of all states affected by COVID-19. (2^nd^ column) Linear regression of the maximum change in park occupancy versus the excess deaths per capita of all states affected by COVID-19. D. (1^st^ column) Linear regression of the initial change in transit occupancy versus the excess deaths per capita of all states affected by COVID-19. (2^nd^ column) Linear regression of the maximum change in transit occupancy versus the excess deaths per capita of all states affected by COVID-19. E. (1^st^ column) Linear regression of the initial change in workplace occupancy versus the excess deaths per capita of all states affected by COVID-19. (2^nd^ column) Linear regression of the maximum change in workplace occupancy versus the excess deaths per capita of all states affected by COVID-19. F. (1^st^ column) Multifactor linear regression of initial change in residential occupancy, maximum change in residential occupancy, population, and deaths. Partial regression plot of initial change in residential occupancy versus excess death. (2^nd^ column) Multifactor linear regression of maximum change in residential occupancy, maximum change in residential occupancy, population, and deaths. Partial regression plot of maximum change in residential occupancy versus excess death. G. Multifactor linear regression of initial change in residential occupancy, maximum change in residential occupancy, population, and deaths. Partial regression plot of state population versus excess death. A-G. Excess deaths are totaled until May 24^th^, 2020. Solid line represents the regression, dashed lines represent 95% confidence intervals. Regression p-values below 0.05 are listed in scientific notation. Adjusted values are generated based on the co-dependencies of all the variables in the model. Red points represent regions that had higher death than predicted by the regression. Grey points represent regions that had lower death than predicted by the regression.

**Supplementary Figure 2:**
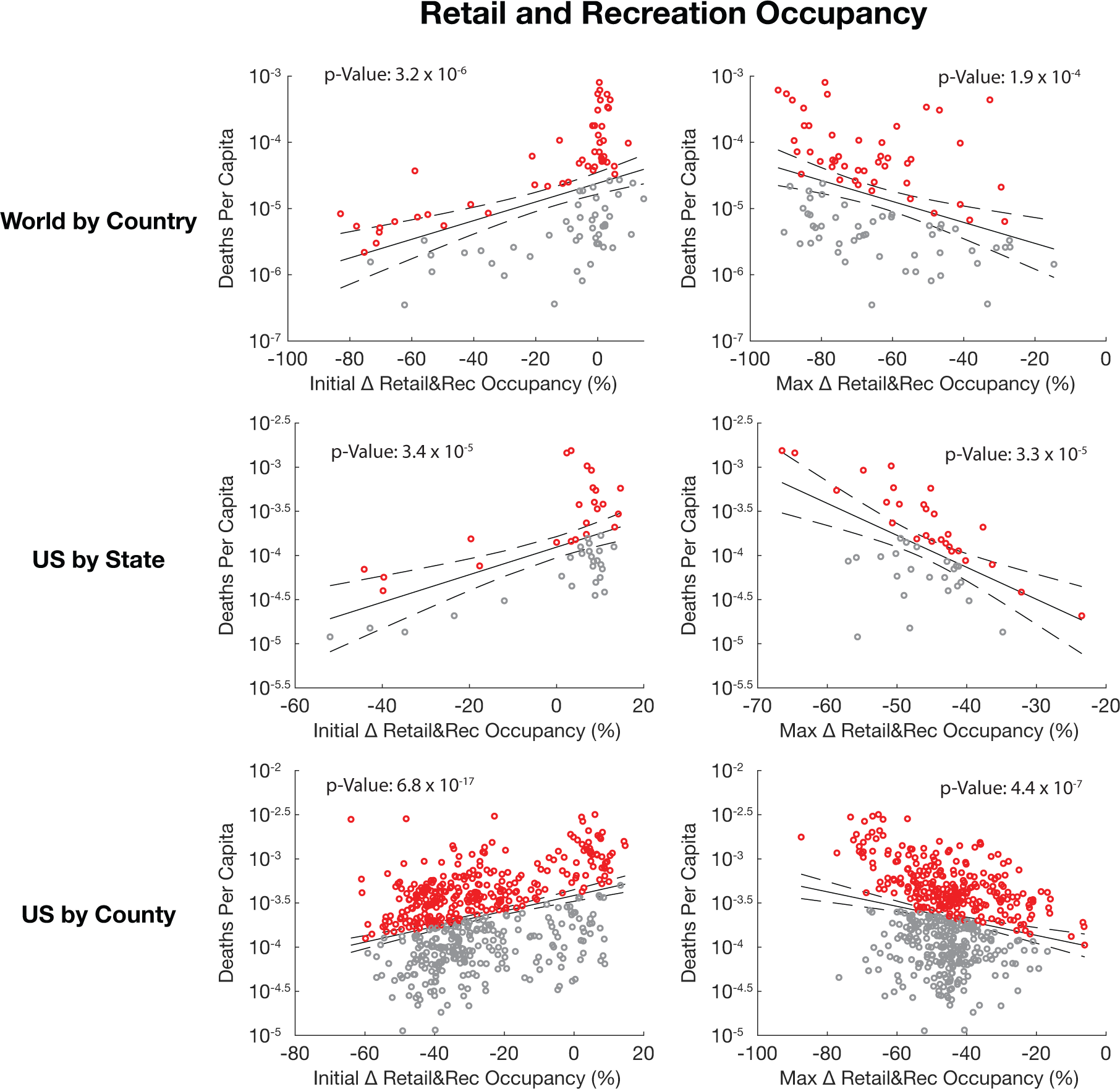
Correlation between recreation occupancy and death also inverts over time. (1^st^ column) Linear regressions of the initial change in retail and recreation occupancy versus the deaths per capita of all regions affected by COVID-19. (2^nd^ column) Linear regressions of the maximum change in retail and recreation occupancy versus the deaths per capita of all regions affected by COVID-19. Rows represent linear regressions done on countries, states, and counties respectively. Deaths per capita are totaled until May 24^th^, 2020. Solid black line represents the linear regression, dashed black lines represent 95% confidence intervals. Regression p-values below 0.05 are listed in scientific notation. Red points represent regions that had higher death than predicted by the regression. Grey points represent regions that had lower death than predicted by the regression.

**Supplementary Figure 3:**
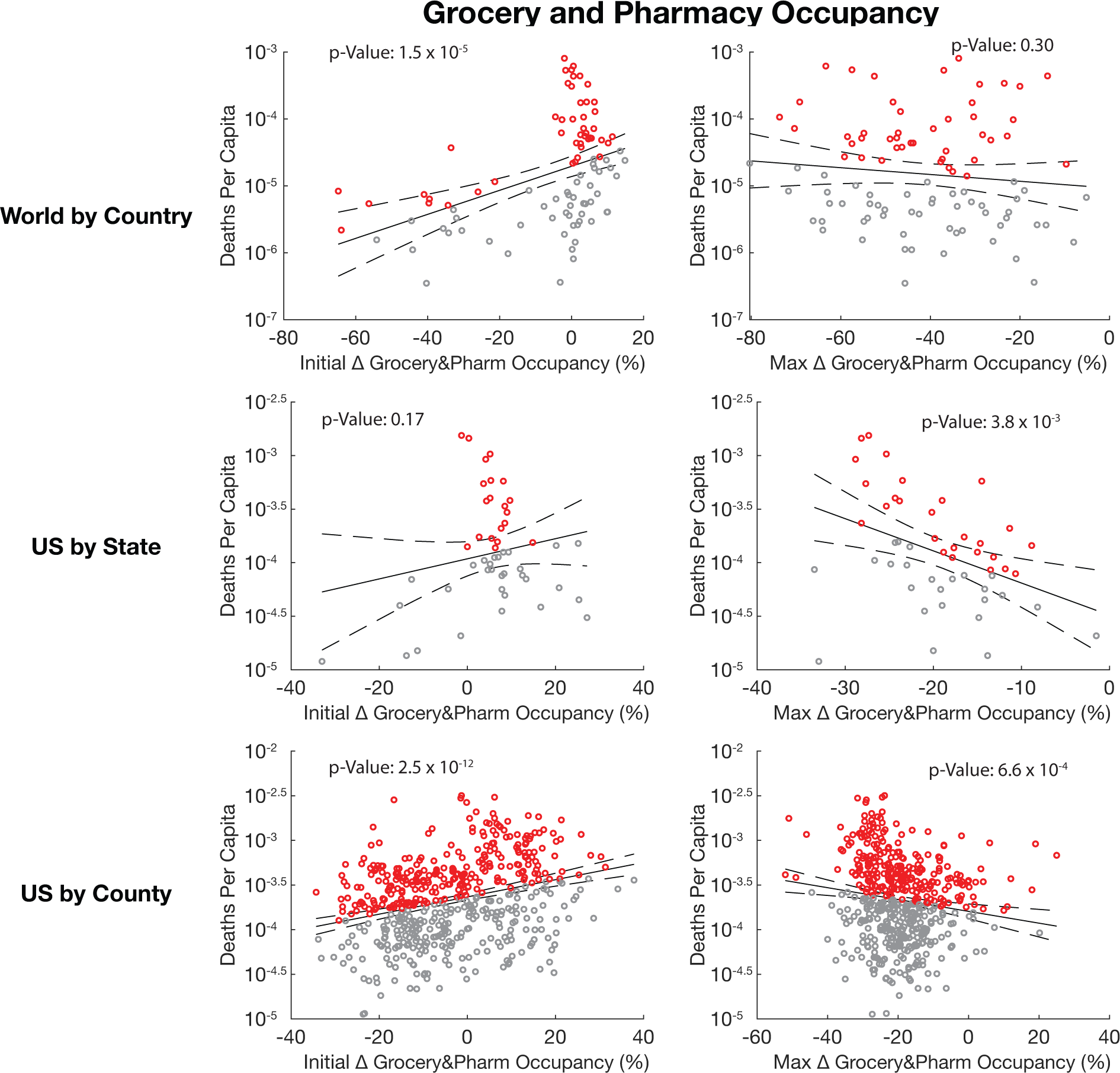
Correlation between grocery occupancy and death also inverts over time. (1^st^ column) Linear regressions of the initial change in grocery and pharmacy occupancy versus the deaths per capita of all regions affected by COVID-19. (2^nd^ column) Linear regressions of the maximum change in grocery and pharmacy occupancy versus the deaths per capita of all regions affected by COVID-19. Rows represent linear regressions done on countries, states, and counties respectively. Deaths per capita are totaled until May 24^th^, 2020. Solid black line represents the linear regression, dashed black lines represent 95% confidence intervals. Regression p-values below 0.05 are listed in scientific notation. Red points represent regions that had higher death than predicted by the regression. Grey points represent regions that had lower death than predicted by the regression.

**Supplementary Figure 4:**
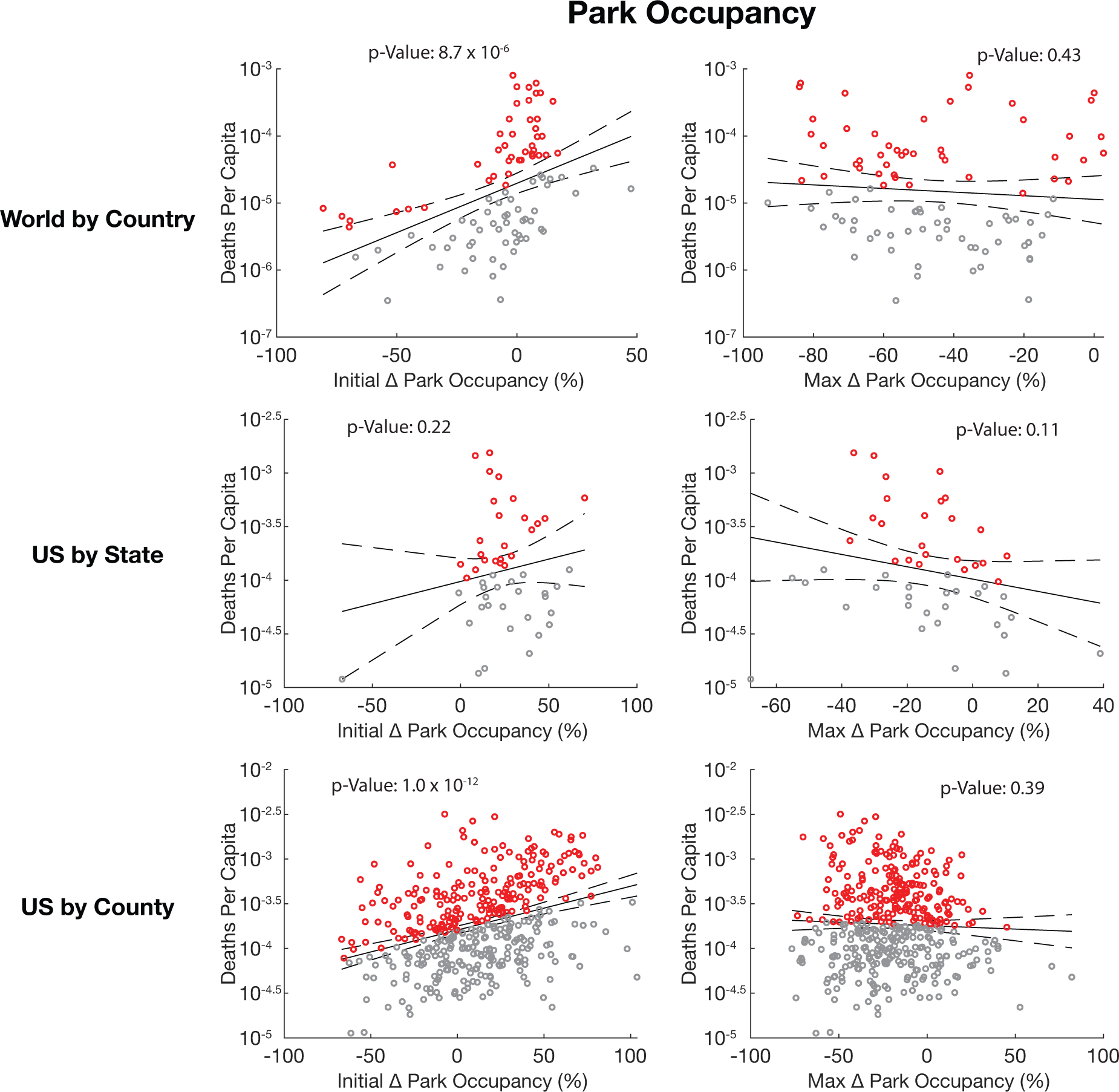
Correlation between park occupancy categories and death does not invert over time. (1^st^ column) Linear regressions of the initial change in park occupancy versus the deaths per capita of all regions affected by COVID-19. (2^nd^ column) Linear regressions of the maximum change in park occupancy versus the deaths per capita of all regions affected by COVID-19. Rows represent linear regressions done on countries, states, and counties respectively. Deaths per capita are totaled until May 24^th^, 2020. Solid black line represents the linear regression, dashed black lines represent 95% confidence intervals. Regression p-values below 0.05 are listed in scientific notation. Red points represent regions that had higher death than predicted by the regression. Grey points represent regions that had lower death than predicted by the regression.

**Supplementary Figure 5:**
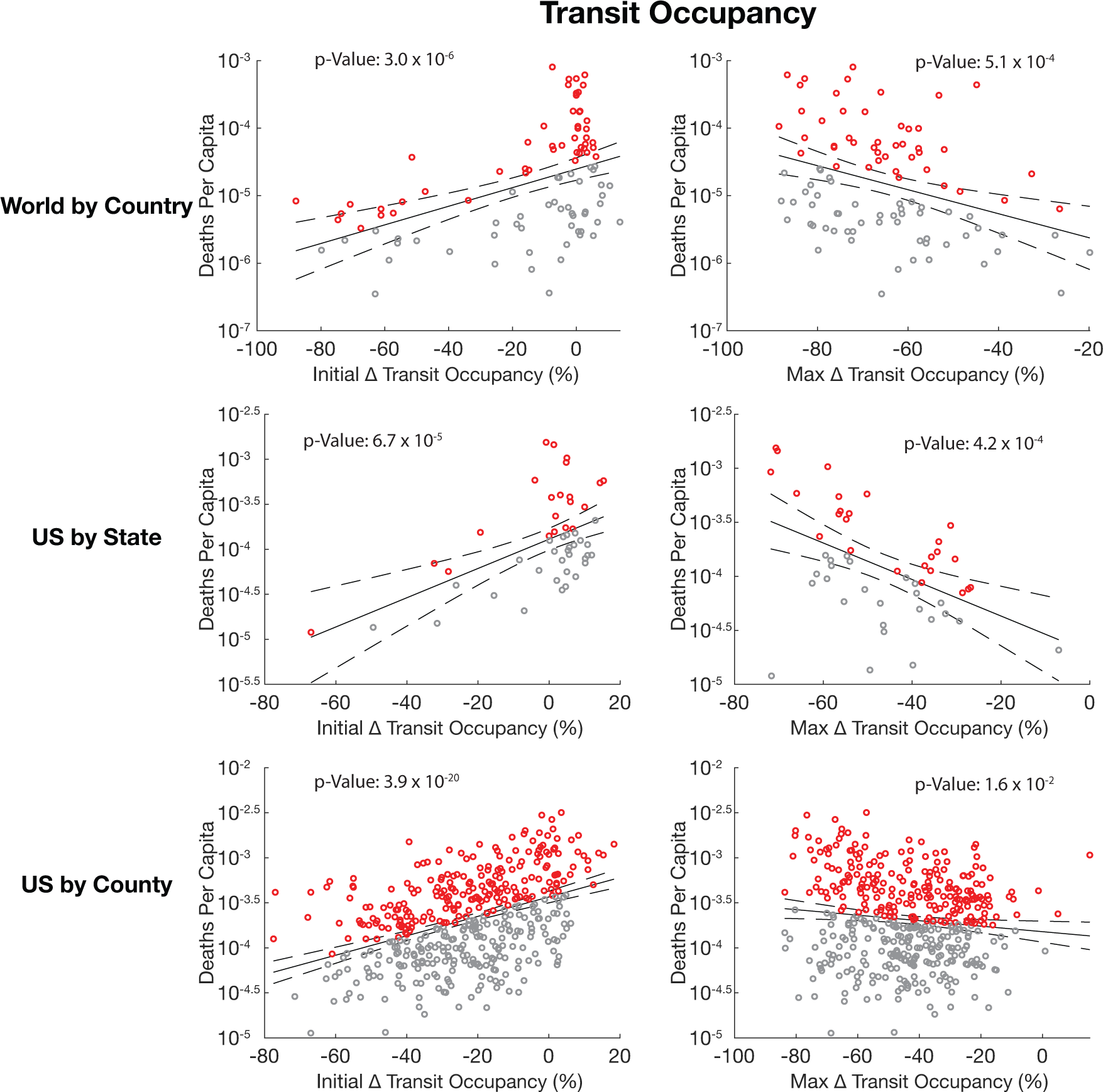
Correlation between transit occupancy and death also inverts over time. (1^st^ column) Linear regressions of the initial change in transit occupancy versus the deaths per capita of all regions affected by COVID-19. (2^nd^ column) Linear regressions of the maximum change in transit occupancy versus the deaths per capita of all regions affected by COVID-19. Rows represent linear regressions done on countries, states, and counties respectively. Deaths per capita are totaled until May 24^th^, 2020. Solid black line represents the linear regression, dashed black lines represent 95% confidence intervals. Regression p-values below 0.05 are listed in scientific notation. Red points represent regions that had higher death than predicted by the regression. Grey points represent regions that had lower death than predicted by the regression.

**Supplementary Figure 6:**
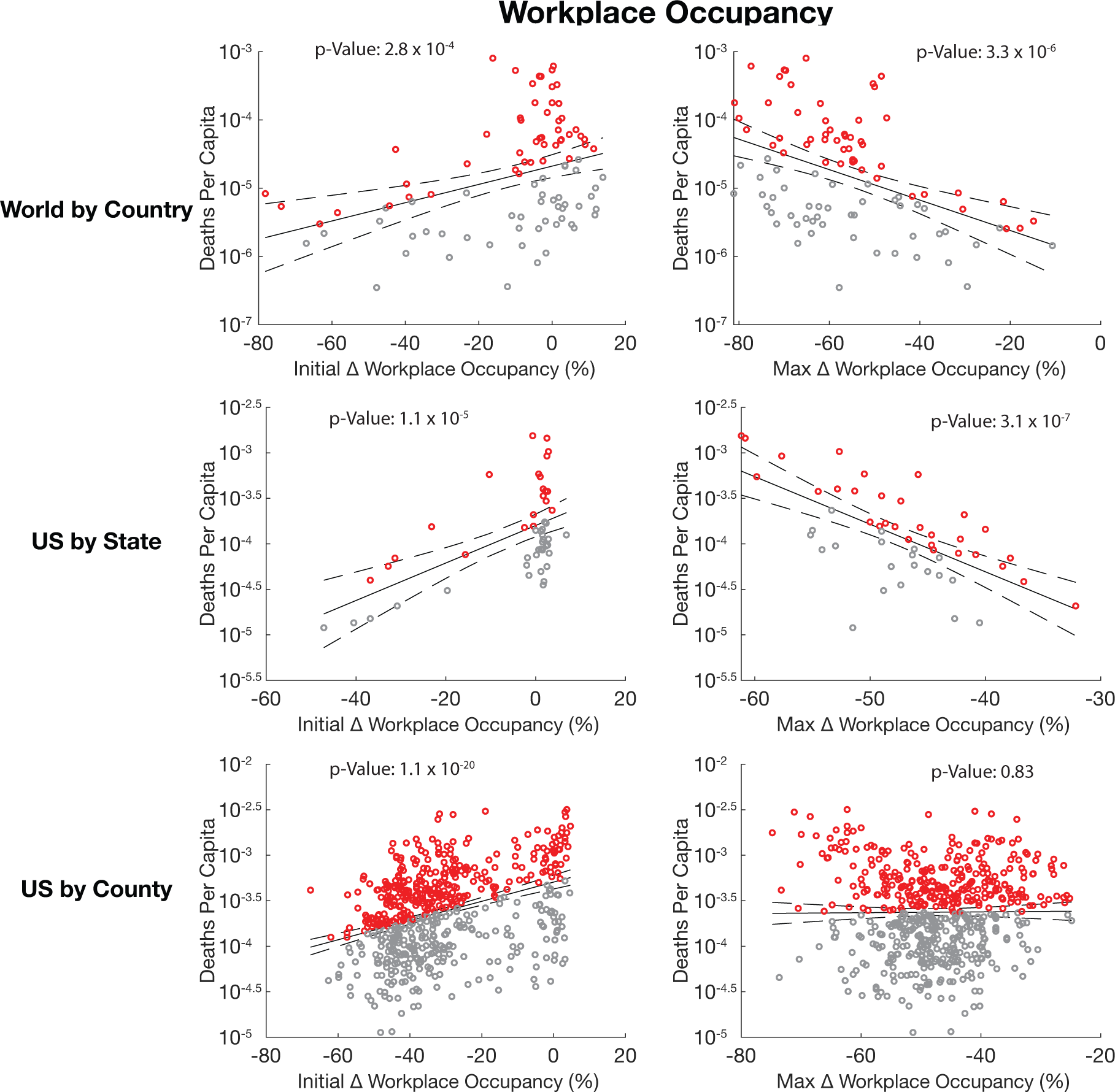
Correlation between workplace occupancy categories and death also inverts over time. (1^st^ column) Linear regressions of the initial change in workplace occupancy versus the deaths per capita of all regions affected by COVID-19. (2^nd^ column) Linear regressions of the maximum change in workplace occupancy versus the deaths per capita of all regions affected by COVID-19. Rows represent linear regressions done on countries, states, and counties respectively. Deaths per capita are totaled until May 24^th^, 2020. Solid black line represents the linear regression, dashed black lines represent 95% confidence intervals. Regression p-values below 0.05 are listed in scientific notation. Red points represent regions that had higher death than predicted by the regression. Grey points represent regions that had lower death than predicted by the regression.

**Supplementary Figure 7:**
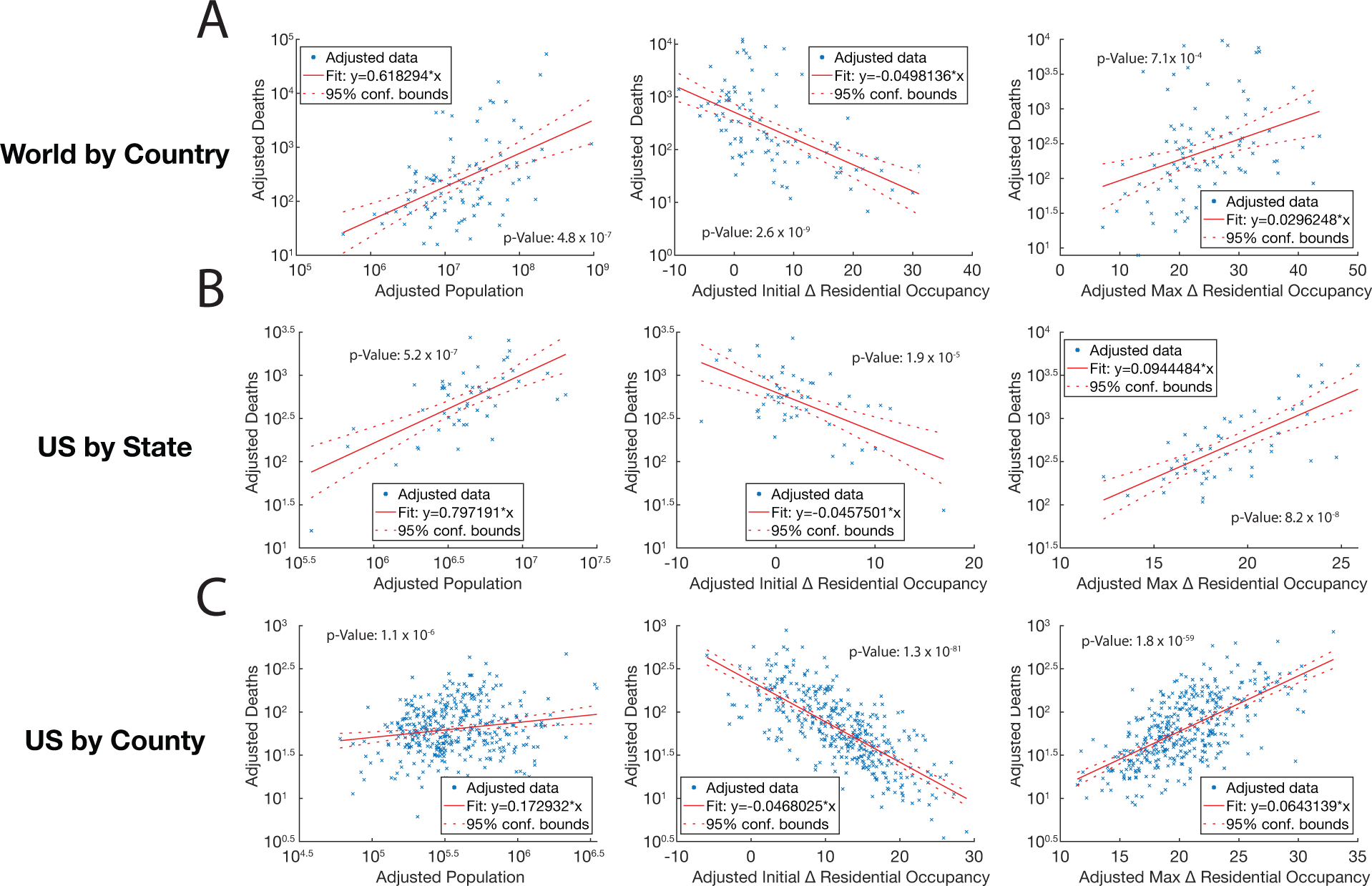
Covariance between occupancy metrics does not cause inverted correlation between residential occupancy and death. A. Multi-factor linear regression between population, initial change in residential occupancy, maximum change in residential occupancy, and confirmed COVID-19 deaths at the country level. (1^st^ column) Partial regression plot of population versus death. (2^nd^ column) Partial regression plot of initial change in residential occupancy versus death. (3^rd^ column) Partial regression plot of max change in residential occupancy versus death. B. Multi-factor linear regression between population, initial change in residential occupancy, maximum change in residential occupancy, and confirmed COVID-19 deaths at the state level. (1^st^ column) Partial regression plot of population versus death. (2^nd^ column) Partial regression plot of initial change in residential occupancy versus death. (3^rd^ column) Partial regression plot of max change in residential occupancy versus death. C. Multi-factor linear regression between population, initial change in residential occupancy, maximum change in residential occupancy, and confirmed COVID-19 deaths at the county level. (1^st^ column) Partial regression plot of population versus death. (2^nd^ column) Partial regression plot of initial change in residential occupancy versus death. (3^rd^ column) Partial regression plot of max change in residential occupancy versus death. A-C. Deaths per capita are totaled until May 24^th^, 2020. Solid red line represents the regression, dashed red lines represent 95% confidence intervals. Regression p-values below 0.05 are listed in scientific notation. Adjusted values are generated based on the co-dependencies of all the variables in the model.

**Supplementary Figure 8:**
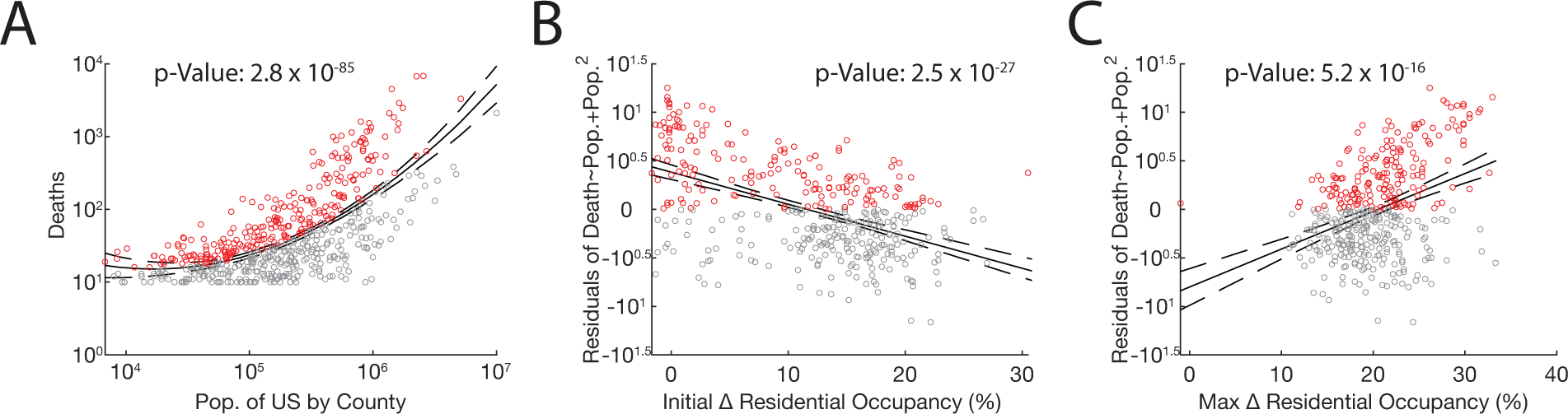
Statistical model of a non-linear correlation between population and death retains inversion of correlation between occupancy and death. A. Quadratic regression between US county population size and the number of confirmed COVID-19 deaths that county experienced as of May 24^th^, 2020. Regression shows that a model that accounts for the effect of population squared on death can capture the nonlinear relationship observed between population and death at the county level. B. Linear regression between the residuals from the quadratic regression in (A) and the initial change in residential occupancy. Regression finds a negative correlation between the two factors, similar to the relationship found in Fig. 1C. C. Linear regression between the residuals from the quadratic regression in (A) and the maximum change in residential occupancy. Regression finds a positive correlation between the two factors, similar to the relationship found in Fig. 1C. A-C. Deaths per capita are totaled until May 24^th^, 2020. Solid black line represents the regression, dashed black lines represent 95% confidence intervals. Regression p-values below 0.05 are listed in scientific notation. Red points represent regions that had higher death than predicted by the quadratic regression. Grey points represent regions that had lower death than predicted by the quadratic regression.

**Supplementary Figure 9:**
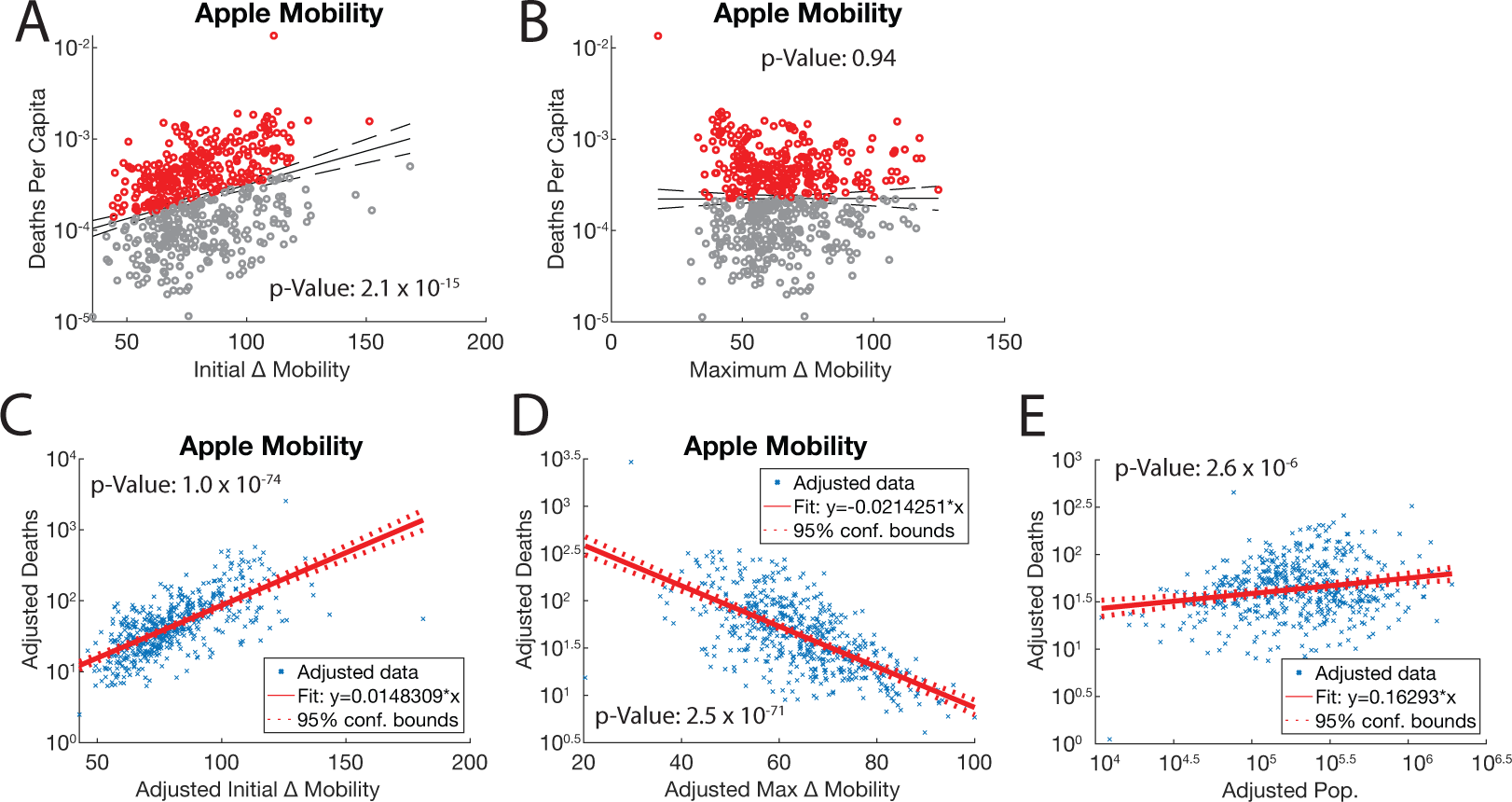
The correlation between Apple mobility and death inverts over time. A. Linear regression of the initial change in driving mobility (from Apple) versus the confirmed deaths per capita of all counties affected by COVID-19. B. Linear regression of the maximum change in driving mobility (from Apple) versus the confirmed deaths per capita of all counties affected by COVID-19. C. Multifactor linear regression of initial change in mobility, maximum change in mobility, population, and deaths. Partial regression plot of initial change in driving mobility (from Apple) versus confirmed deaths at the county level. D. Multifactor linear regression of maximum change in mobility, maximum change in mobility, population, and deaths. Partial regression plot of maximum change in driving mobility (from Apple) versus confirmed deaths at the county level. E. Multifactor linear regression of initial change in mobility, maximum change in mobility, population, and deaths. Partial regression plot of county population versus confirmed deaths. A-E. Confirmed COVID-19 deaths are totaled until May 24^th^, 2020. Solid line represents the regression, dashed lines represent 95% confidence intervals. Regression p-values below 0.05 are listed in scientific notation. Adjusted values are generated based on the co-dependencies of all the variables in the model. Red points represent regions that had higher death than predicted by the regression. Grey points represent regions that had lower death than predicted by the regression. C&D. Linear regression of the first peak of infections per day versus the time it took to reach that peak for real infection counts recorded in countries internationally. Points represent individual regions, solid line represents linear regression, and dotted lines represent 95% confidence interval.

**Supplementary Figure 10:**
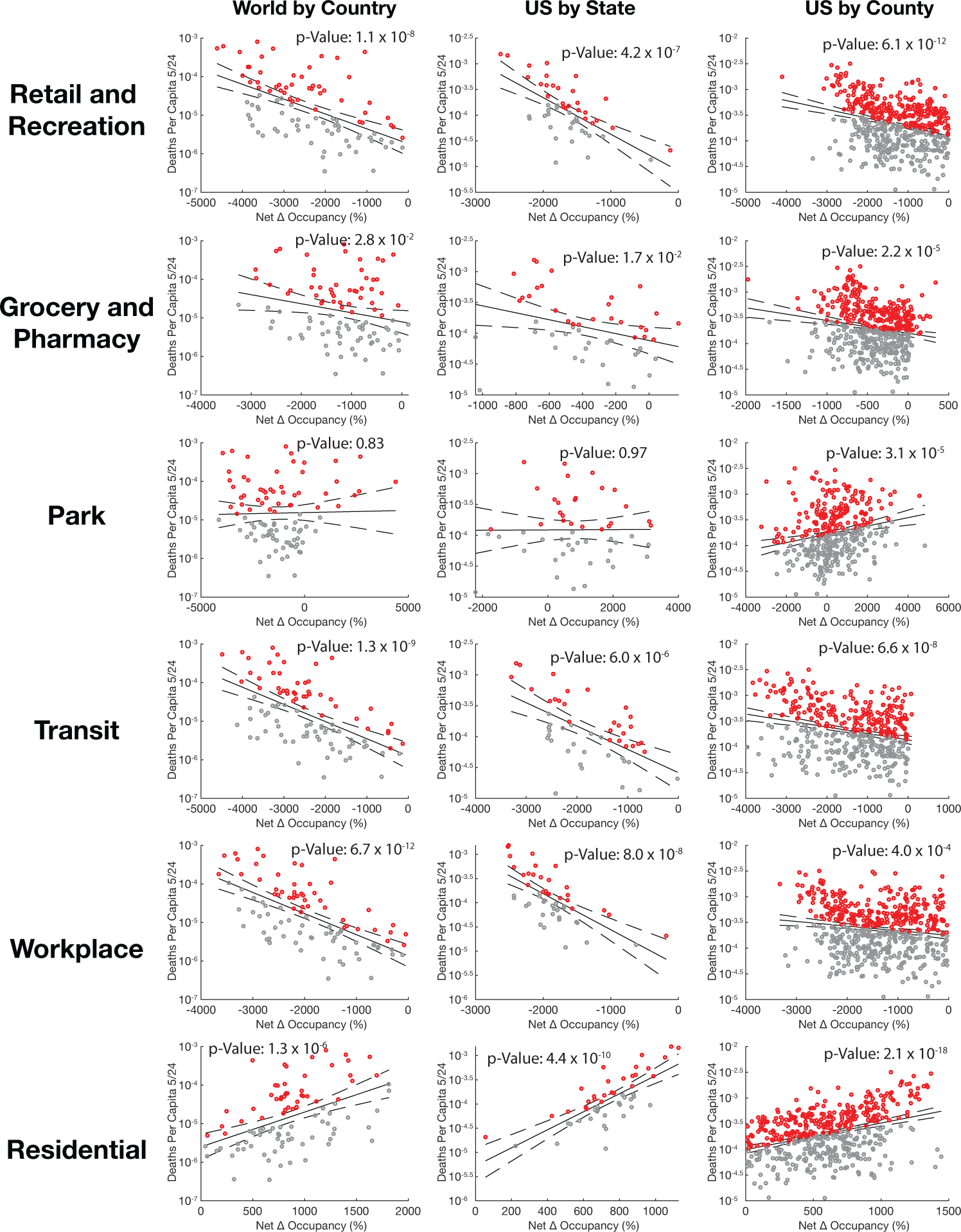
Occupancy under the curve is perversely correlated with COVID-19 mortality. Linear regressions of net changes in residential occupancy versus confirmed deaths per capita. Columns - Country, state, and county data Rows-Retail & recreation, grocery & pharmacy, park, transit, workplace, and residential occupancy. Deaths per capita are totaled until May 24^th^, 2020. Solid black line represents the linear regression, dashed black lines represent 95% confidence intervals. Regression p-values below 0.05 are listed in scientific notation. Red points represent regions that had higher death than predicted by the regression. Grey points represent regions that had lower death than predicted by the regression.

**Supplementary Figure 11:**
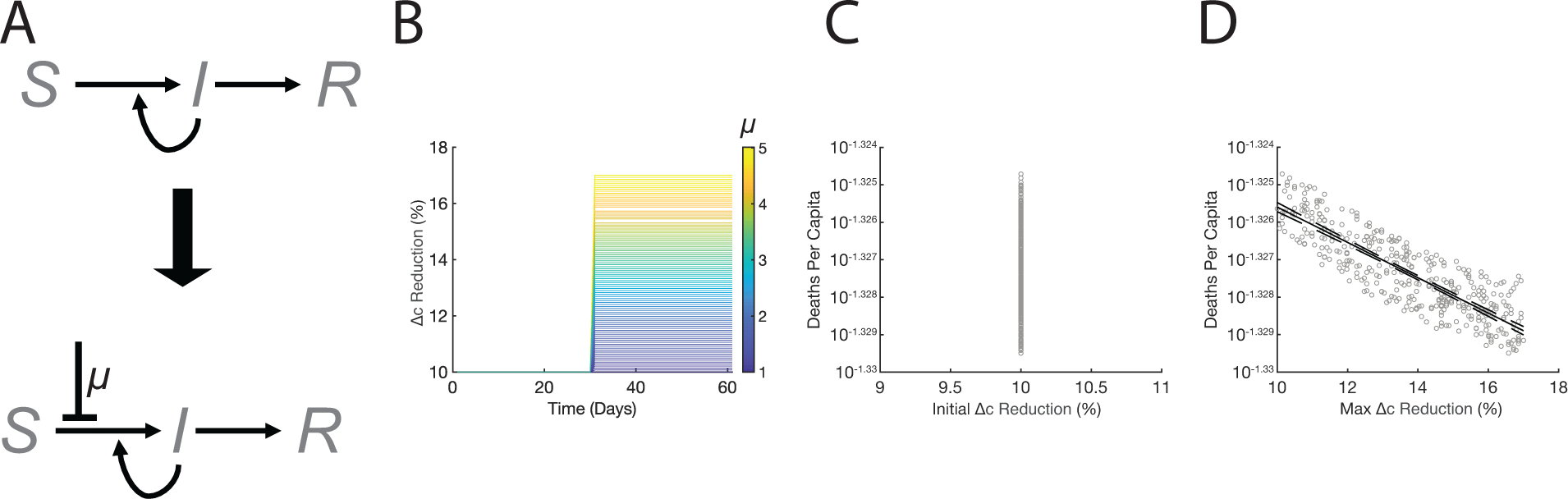
Dynamic changes in induced sequestration are not sufficient to generate inverted occupancy-death correlation. A. Schematic of epidemic ODE model with induced sequestration implemented partway through epidemic. B. Trends in contact rate reduction over time as determined by delayed induced sequestration. Stronger induced sequestration values (*μ*) lead to a larger reduction in contact rates. Induced sequestration is applied after 30 days in this simulation and remains constant for 30 days after application. C. Linear regression of simulated deaths versus initial changes in contact rate reduction generated by an SIR model with policy implemented partway through epidemic. Initial changes in contact rate reduction are not correlated with any changes in death. D. Linear regression of simulated deaths versus maximum changes in contact rate reduction generated by an SIR model with induced sequestration implemented partway through epidemic. Maximum changes in contact rate reduction are negatively correlated with death (p-value: 8.9 × 10^−110^).

**Supplementary Figure 12:**
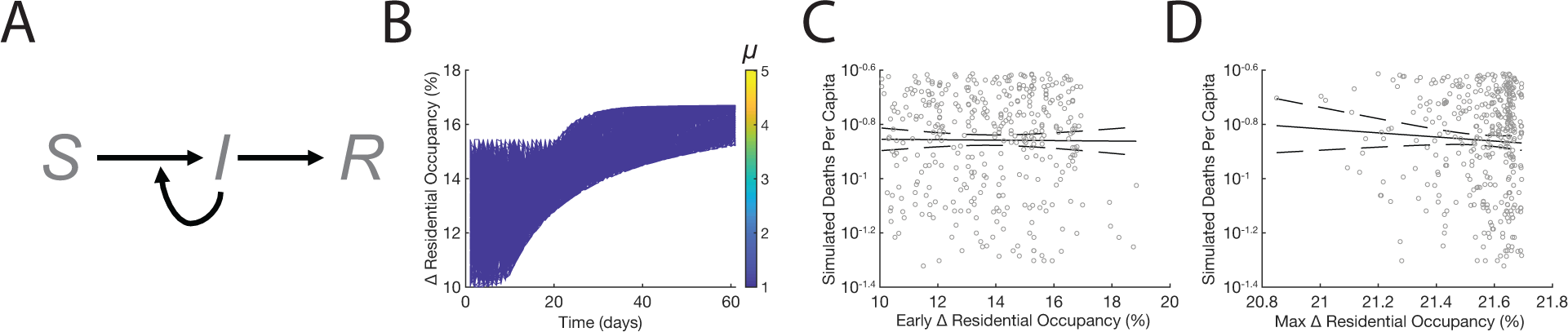
Coincidental population sequestration during an epidemic is not sufficient to invert mobility-death correlation. A. Schematic of the simple SIRD model [**Eqs. 1-4**] with no sequestration [**Eq. S1**] where contact-reduction rate c = 1. B. Example curves of randomly selected residential occupancy values taken from an increasing distribution. This represents a scenario where residential occupancy increases coincidentally with the emergence of COVID-19, but has no functional relationship with the pandemic. C-D. Numerical simulations of mortality (calculated as 1% of R/N) as a function of initial and max changes in simulated residential occupancy from simple SIR model (regression p-values=0.76 and 0.99).

**Supplementary Figure 13:**
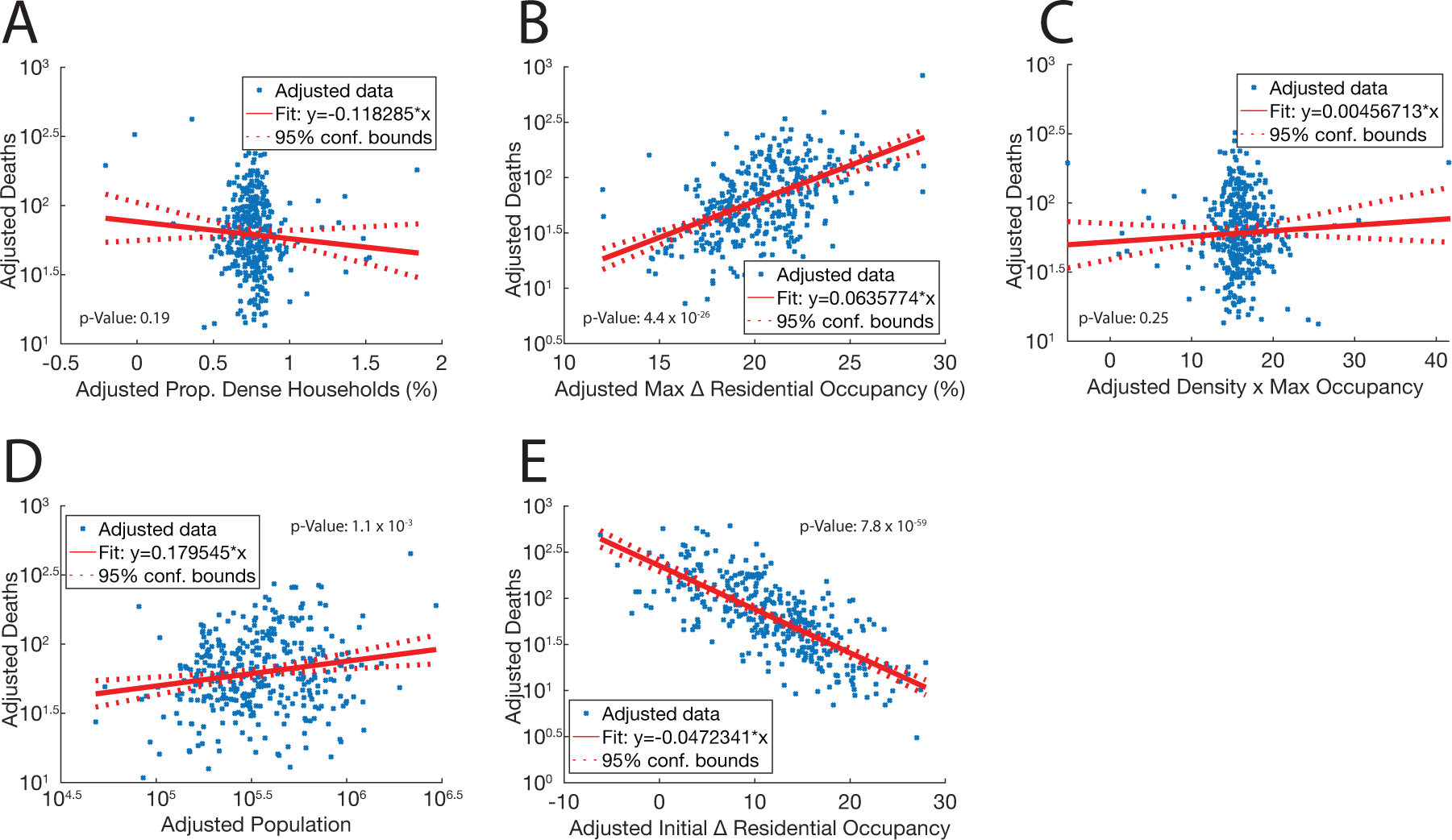
Density and maximum residential occupancy do not have a synergistic correlation with death. Multi-factor linear regression between proportion of dense households, maximum change in residential occupancy, interaction between household density and maximum residential occupancy, population, initial change in residential occupancy, and confirmed COVID-19 deaths at the county level. A. Partial regression plot of household density proportion as measured by the percentage of homes in a county that had more than 1.5 people per room. B. Partial regression plot of max change in residential occupancy versus death. C. Partial regression plot of interaction between household density and maximum occupancy. D. Partial regression plot of population versus death. E. Partial regression plot of initial change in residential occupancy versus death. A-E. Deaths per capita are totaled until May 24^th^, 2020. Solid red line represents the regression, dashed red lines represent 95% confidence intervals. Regression p-values below 0.05 are listed in scientific notation. Adjusted values are generated based on the co-dependencies of all the variables in the model.

**Supplementary Figure 14:**
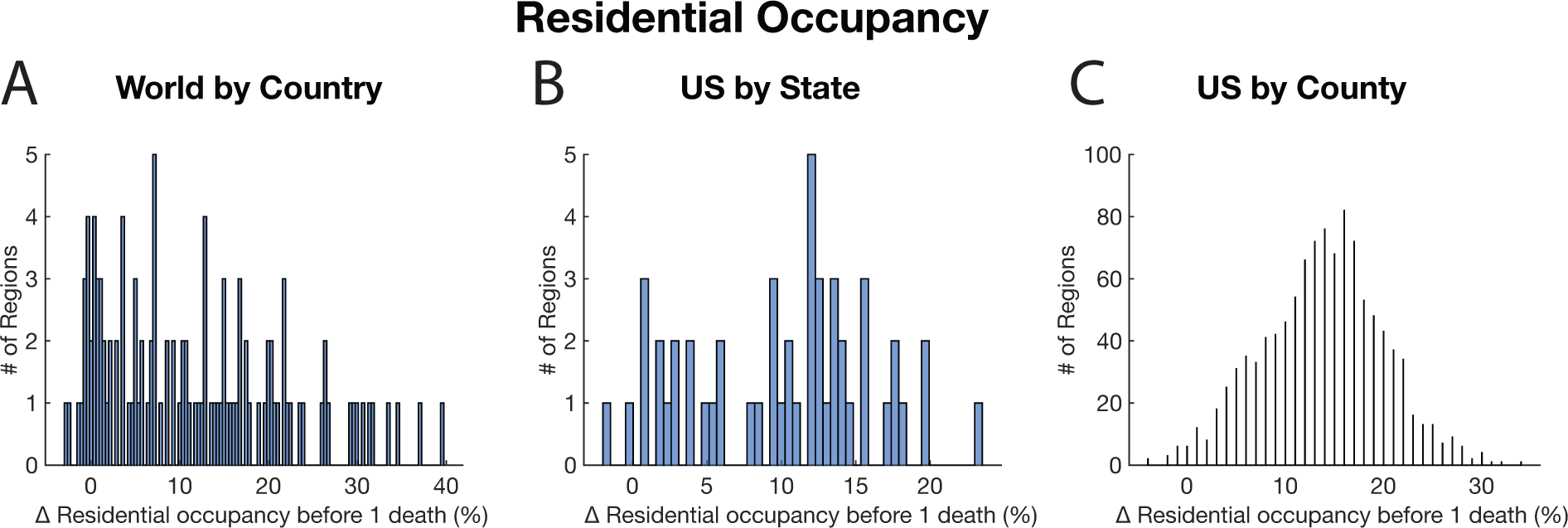
Changes in residential occupancy occur before local deaths are recorded. A. Histogram of the change in residential occupancy that occurs in countries before one death is recorded in that country. B. Histogram of the change in residential occupancy that occurs in states before one death is recorded in that state. C. Histogram of the change in residential occupancy that occurs in counties before one death is recorded in that county.

**Supplementary Figure 15:**
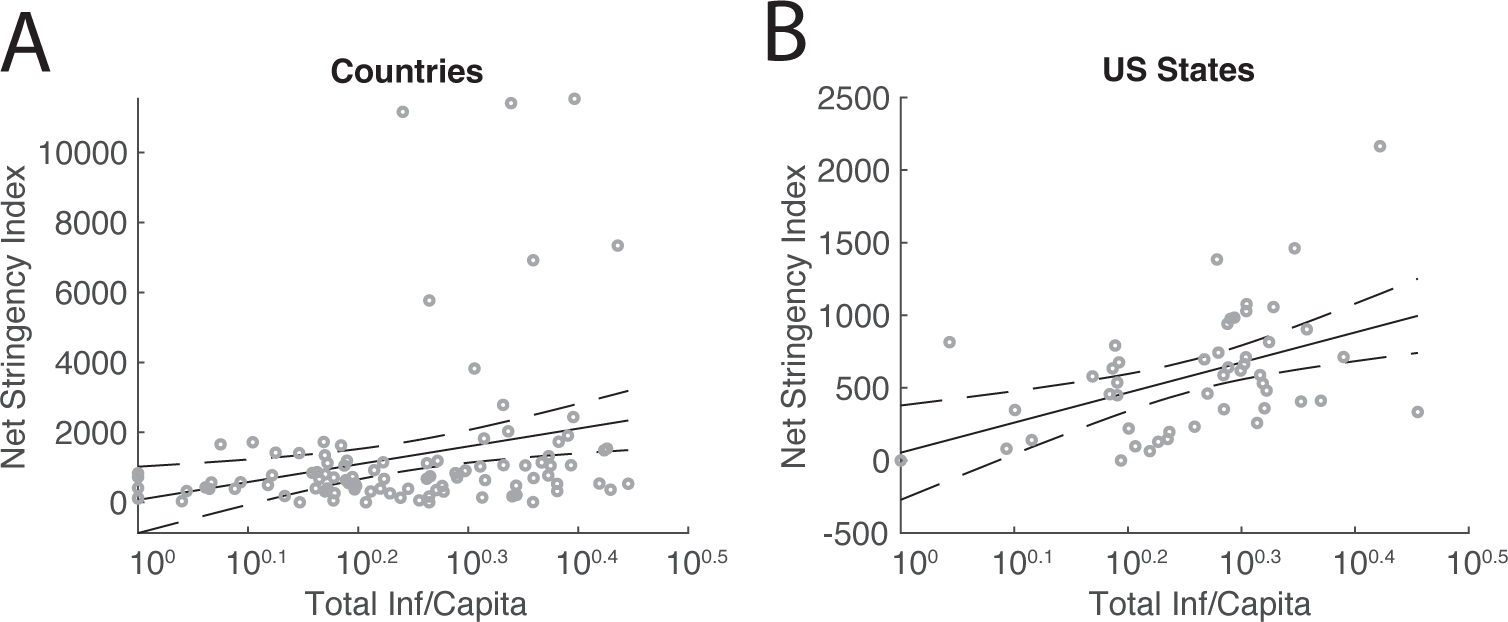
Early changes in policy are correlated with infection count. A. Regression of total infections per capita versus net stringency index for countries affected by COVID-19. Infections per capita were totaled before a single death occurred in the region, and the net stringency index was calculated over the same time period. The p-value of the linear regression = 5.8 × 10^−3^ (dashed lines represent 95% confidence interval). B. Regression of total infections per capita versus net stringency index for US states affected by COVID-19. Infections per capita were totaled before a single death occurred in the region, and the net stringency index was calculated over the same time period. The p-value of the linear regression = 9.6 × 10^−4^ (dashed lines represent 95% confidence interval).

**Supplementary Figure 16:**
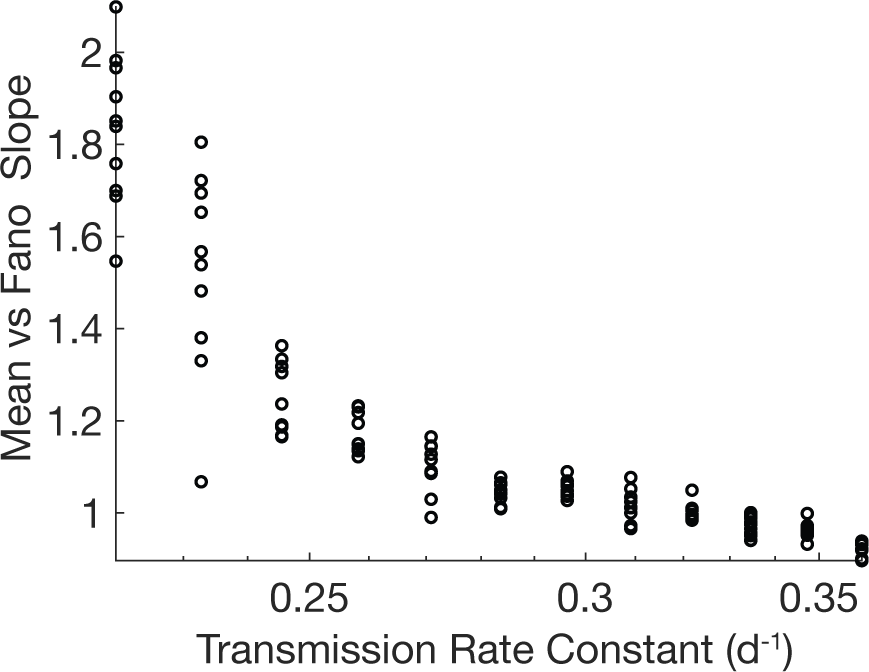
Modulation of transmission rate constant is not sufficient to control infection rate variation. Summary diagram of the effect that changing the transmission rate constant (*β*) has on the regressed slope of simulated Fano factor versus mean plots. Graph shows that increasing the transmission rate constant can reduce the slope to one, but not below one.

**Supplementary Figure 17:**
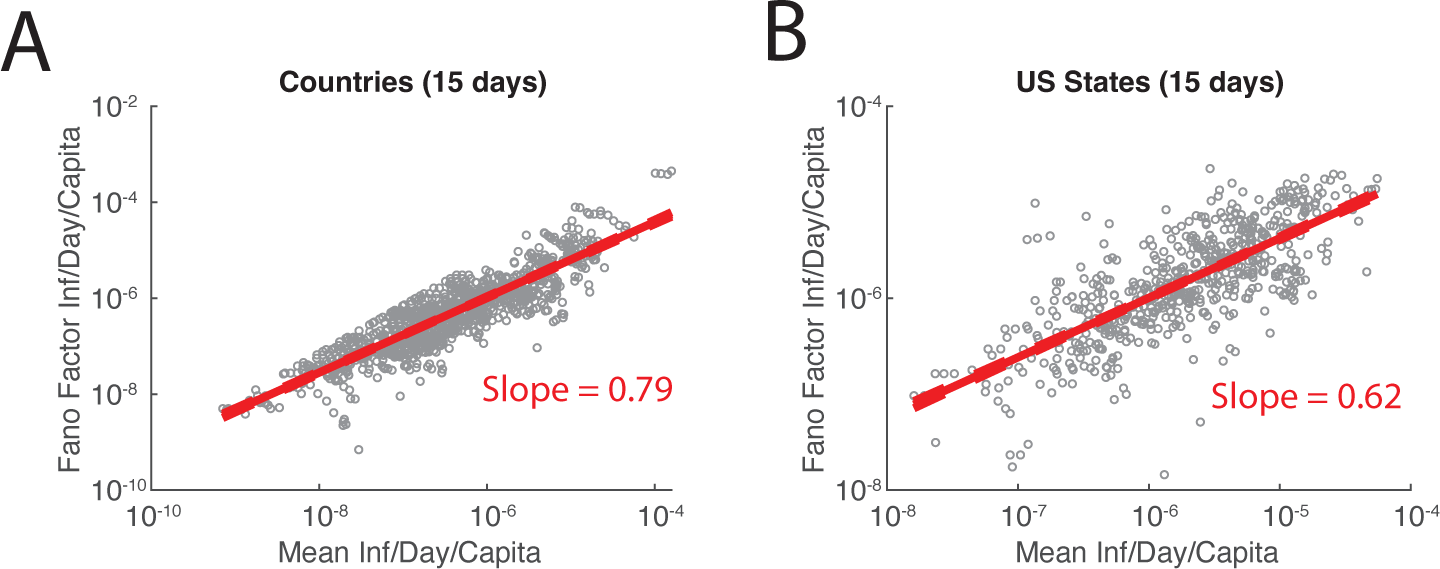
Early variation in infections per day is suppressed. A. Data of Fano factor versus mean of the first 15 days of infection for countries affected by COVID-19. Slope of linear regression = 0.79 (dashed red lines represent 95% confidence interval). B. Fano factor versus mean plot of the first 15 days of infection for US states affected by COVID-19. Linear regression slope = 0.62 (dashed red lines represent 95% confidence interval).

**Supplementary Figure 18:**
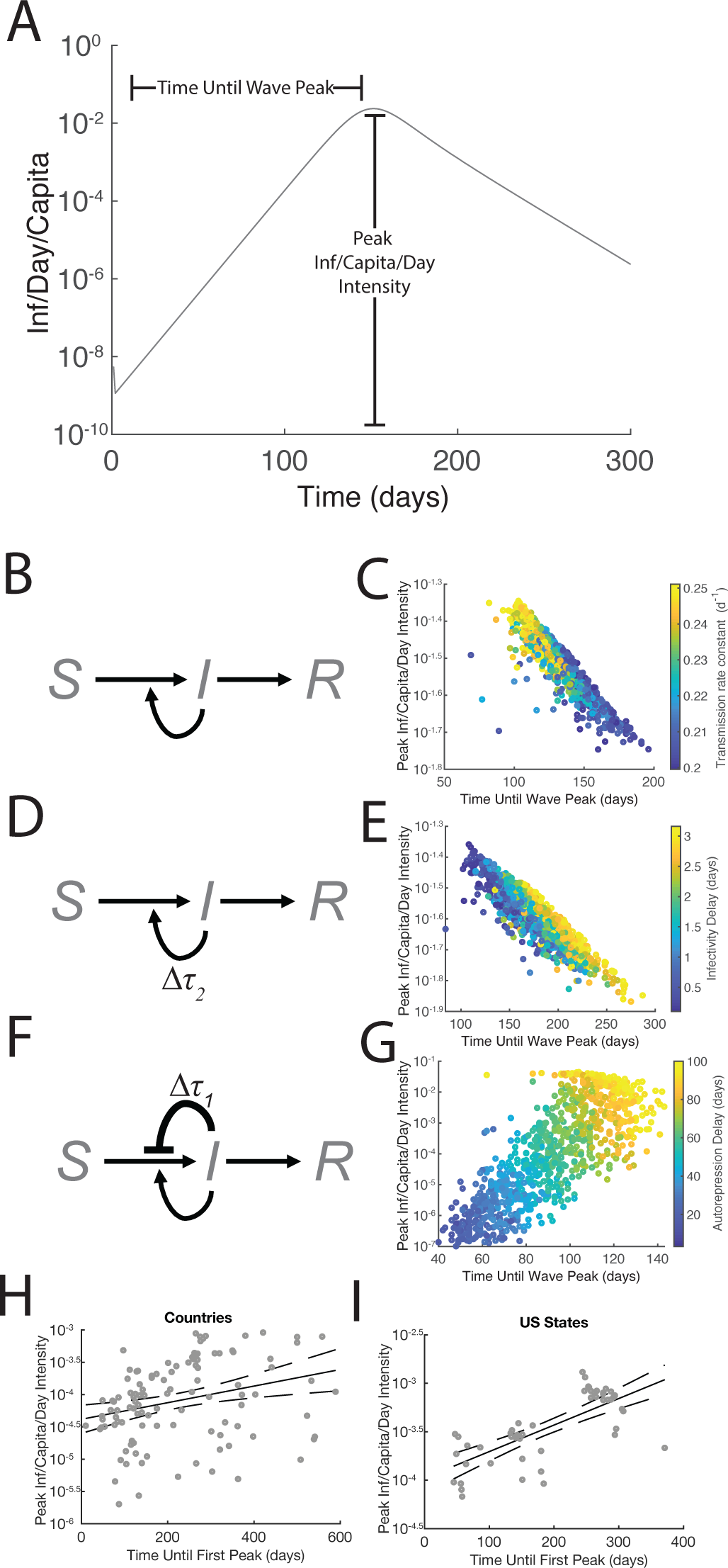
Strengthening infectivity leads to stronger, faster waves. A. Example simulated epidemic to illustrate the calculation of parameters necessary to quantify the timing and intensity of infection waves. B. Schematic of the simple SIR model [**Eqs. 1-4**]. C. Plot of a series of epidemic simulations where infection wave timing vs intensity has been quantified. Color of points represents value of transmission rate constant (*β*). D. Schematic of the simple SIR model with a delay in susceptible individuals becoming infective [**Eqs. S3-S6**]. E. Plot of a series of epidemic simulations where infection wave timing vs intensity has been quantified. Color of points represents value of infectivity delay (*τ*_2_). F. Schematic of the SIR model with delayed autorepression [**Eqs. 1-4, S2**]. G. Plot of a series of epidemic simulations where infection wave timing vs intensity has been quantified. Color of points represents value of autorepression delay (*τ*_1_). H. Linear regression of measured values for time until first peak of infections per day versus peak infections per day at the international scale, p-value: 2.5 × 10^−3^. Points represent individual regions, solid line represents linear regression, and dotted lines represent 95% confidence interval. I. Linear regression of measured values for time until first peak of infections per day versus peak infections per day at the US state scale, p-value: 2.7 × 10^−9^. Points represent individual regions, solid line represents linear regression, and dotted lines represent 95% confidence interval.

